# AI-enabled proteomic profiling identifies interpretable diagnostic biomarkers for gastric-type adenocarcinoma of the uterine cervix

**DOI:** 10.64898/2026.06.24.26356417

**Authors:** Tao Zhu, Han Huang, Zhaoxu Yang, Maowei Ni, Yun Xi, Yulan Liu, Wenjie Luo, Xuedong Tang, Xiaomei Fan, Chen Yang, Zhen Shen, Jincheng Wang, Huarong Tang, Qinjie Weng

**Affiliations:** Center for Drug Safety Evaluation and Research, Zhejiang Province Key Laboratory of Anti-Cancer Drug Research, School of Pharmacy, Zhejiang University, Hangzhou, Zhejiang 310058, China; Department of Gynecological Oncology, Zhejiang Cancer Hospital, Hangzhou, Zhejiang 310022, China; Taizhou Institute of Zhejiang University, Taizhou, Zhejiang 318000, China; Zhejiang Cancer Hospital, Hangzhou Institute of Medicine (HIM), Chinese Academy of Sciences, Hangzhou, Zhejiang 310022, China; Department of Pathology, Zhejiang Cancer Hospital, Hangzhou, Zhejiang 310022, China; ZJU-Xinchang Joint Innovation Center (TianMu Laboratory), Xinchang, Zhejiang 312500, China; Department of Gynecology, Jiaxing Maternity and Child Health Care Hospital, Jiaxing, Zhejiang 314000, China; The Fourth Hospital of Hebei Medical University, Shijiazhuang, Hebei 050000, China; Department of Obstetrics and Gynecology, The Second Affiliated Hospital of Anhui Medical University, Hefei, Anhui 230601, China; Department of Gynecological Radiotherapy, Zhejiang Cancer Hospital, Hangzhou Institute of Medicine (HIM), Chinese Academy of Sciences, Hangzhou, Zhejiang 310022, China; The Second Affiliated Hospital, Zhejiang University School of Medicine, Hangzhou, Zhejiang 310009, China; Institute of Fundamental and Transdisciplinary Research, Zhejiang University, Hangzhou, Zhejiang 310058, China

## Abstract

Gastric-type adenocarcinoma of the uterine cervix (GAS) is a rare, aggressive cervical cancer unrelated to HPV infection that is frequently misdiagnosed because it closely resembles both benign cervical glands and HPV-related cervical adenocarcinoma. This diagnostic confusion can lead to inappropriate treatment, highlighting the need for objective molecular markers. Here, we performed the systematic multi-center proteomic analysis of GAS, profiling 407 cervical tissue samples to map its molecular landscape. To overcome limited sample size and biological noise, we developed WEDGE. First, generative AI synthesizes realistic artificial proteomic profiles to augment the training data. Biologically informed network analysis then leverages known biological relationships to surface diagnostically meaningful patterns. WEDGE identified a two-protein signature, Pepsinogen C (PGC) and DNA Methyltransferase 1 (DNMT1), that distinguished GAS from HPV-related cervical cancer with 93% accuracy in the test cohort and 97% accuracy in an external proteomic cohort, outperforming existing biomarker-discovery methods. Tissue staining of an IHC validation cohort confirmed the expression patterns and reached a diagnostic accuracy of 87.9%. Beyond diagnosis, PGC independently predicted patient outcomes, and combining PGC with routine clinical features improved risk prediction (C-index 0.701). Together, these results establish an AI-driven framework for biomarker discovery and provide clinically relevant candidate tools for diagnosing and prognosticating for GAS.

**Teaser:** A graph-based proteomic framework prioritizes a two-protein IHC-amenable panel for gastric-type cervical adenocarcinoma.

## INTRODUCTION

Cervical adenocarcinoma (CA) represents a biologically and clinically heterogeneous group of malignancies(*1*), encompassing both human papillomavirus (HPV)-associated and HPV-independent subtypes(*2*). While the incidence of HPV-driven cancers is declining due to vaccination and screening programs, HPV-independent subtypes represent a rising clinical challenge(*3*). Among them, gastric-type adenocarcinoma of the uterine cervix (GAS) is rare but accounts for the majority of cases within this category(*4*). GAS exhibits highly aggressive invasion and metastatic behavior and markedly poor prognosis compared with HPV-associated adenocarcinoma (HPV-CA)(*5*). Therefore, precise diagnosis for GAS is crucial for appropriate therapeutic decision-making and prognostic assessment.

Despite its clinical significance, GAS remains difficult to diagnose precisely in routine practice(*6*). First, highly differentiated forms of GAS, such as minimal deviation adenocarcinoma (MDA), often mimic normal glands so closely that pathologists easily mistake them for benign tissue(*7*). Furthermore, some adenocarcinomas exhibit a hybrid appearance suggestive of both HPV-CA and GAS, even though these two are etiologically distinct entities rather than true mixed tumors. Consequently, this overlapping morphology often leads to misclassification in daily practice. (*8*). Conventional screening modalities, including high-risk HPV DNA testing, Thin-prep Cytologic Test (TCT), and colposcopy(*9*), often fail to detect GAS due to its atypical clinical presentation and HPV-independent nature. These diagnostic pitfalls, coupled with the aggressive nature of GAS, underscore an urgent need for novel and reliable biomarkers with high sensitivity and specificity.

Proteomics has recently emerged as a powerful technology in cancer research(*10*), particularly for discovery of diagnostic biomarkers(*11–13*). By enabling systematic and large-scale profiling of protein expression, proteomics provides a comprehensive overview that integrates both genotypic and phenotypic alterations of tumors(*14*), including dynamic changes during tumor progression and within tumor microenvironment(*15, 16*). Such protein-level alterations often reflect disease biology more directly than genomic or transcriptomic approaches. However, GAS has not yet been characterized through systematic, cohort-based proteomics. This critical gap severely limits the discovery of specific diagnostic and prognostic biomarkers to guide personalized treatment strategies and improve clinical outcomes for GAS patients.

Conventional proteomic biomarker discovery approaches predominantly rely on statistical testing to identify differentially expressed proteins(*17–19*), treating molecules largely in isolation and often neglecting the complex biological networks that drive tumor progression. These limitations are further exacerbated in rare malignancies like GAS, where restricted cohort sizes severely compromise statistical power and make clinical utility of biomarkers identified from such underpowered cohorts(*20, 21*). Although deep learning (DL) methods offer powerful capabilities for modeling high-dimensional data and capturing nonlinear relationships(*22*), their application in rare diseases is frequently hindered by the inherent scarcity of training data. Moreover, traditional data augmentation strategies commonly used in imaging domains are not readily applicable to proteomic data, where features are governed by intricate biological dependencies rather than spatial structure(*23*).

To address these dual challenges of biological obscurity and data sparsity, we present a systematic proteomic characterization of GAS derived from a multi-center cohort of 407 participants. Beyond constructing a proteomic landscape, we developed a deep learning framework termed WGAN-GP Enhanced Dual-stream GCN for Expression-Matrix (WEDGE). This methodological innovation integrates Wasserstein Generative Adversarial Networks (WGAN-GP)(*24*) with Dual-stream Graph Convolutional Networks (GCN) to overcome data scarcity. Unlike the transcriptomics-oriented GAN-based augmentation methods of MDWGAN-GP(*25*) and omicsGAN(*26*), WEDGE is designed for tabular proteomic matrices and prioritizes a graph-aware downstream classifier. Specifically, WGAN-GP generates high-fidelity synthetic proteomic profiles, while the dual-stream GCN incorporates prior biological knowledge of protein-protein interactions and gene regulatory networks.

Here, we demonstrate that WEDGE significantly outperforms traditional statistical and machine learning approaches in identifying diagnostic markers. Utilizing this framework, we identified and validated a two-protein diagnostic panel (PGC and DNMT1) that exhibited superior performance in distinguishing GAS from HPV-CA and demonstrated its clinical translatability through independent immunohistochemical validation.

Furthermore, we highlighted PGC as a critical prognostic indicator and constructed a risk stratification model that effectively improves outcome prediction. Collectively, our work provides both a clinically actionable biomarker set and a generalizable AI-driven proteomic framework for biomarker discovery in rare malignancies.

## RESULTS

### Proteomic landscape and molecular characterization of GAS

To establish a comprehensive proteomic landscape of cervical malignancies and identify robust markers for gastric-type adenocarcinoma of the uterine cervix (GAS), we assembled a multi-center tissue repository encompassing 407 specimens (Fig. 1A). The initial baseline cohort spanned four distinct histological categories: 125 specimens with GAS, 136 with HPV-associated cervical adenocarcinoma (HPV-CA), 16 with rare non-GAS HPV-independent variants (11 clear cell and 5 mesonephric adenocarcinomas), and 130 morphologically normal cervical tissues serving as controls. To evaluate geographical boundaries and center-specific variations, these samples were strategically derived from two clinical institutions: Zhejiang Cohort contained 378 specimens harvested from Zhejiang Cancer Hospital, and Jiaxing Cohort comprised 29 independent specimens from Jiaxing Maternity and Child Health Care Hospital. For downstream diagnostic modeling, we enforced a rigorous filtration workflow to isolate disease-specific signals: we confined the 130 normal controls (Con) and 16 rare non-GAS variants strictly to initial descriptive landscape mapping and subsequently excluded them from the predictive pipeline. We partitioned the remaining 261 disease-derived specimens (125 GAS and 136 HPV-CA) to test center-level generalizability. Specifically, the 232 disease specimens from Zhejiang Cohort formed the Development Cohort, which was further stratified into a Training Cohort (n = 144; 73 GAS and 71 HPV-CA) and an Internal Test Cohort (n = 88; 41 GAS and 47 HPV-CA). Concurrently, we assigned the 29 disease specimens from Jiaxing Cohort as an independent, external cohort to assess cross-center performance (Fig. 1A).

**Fig. 1.**
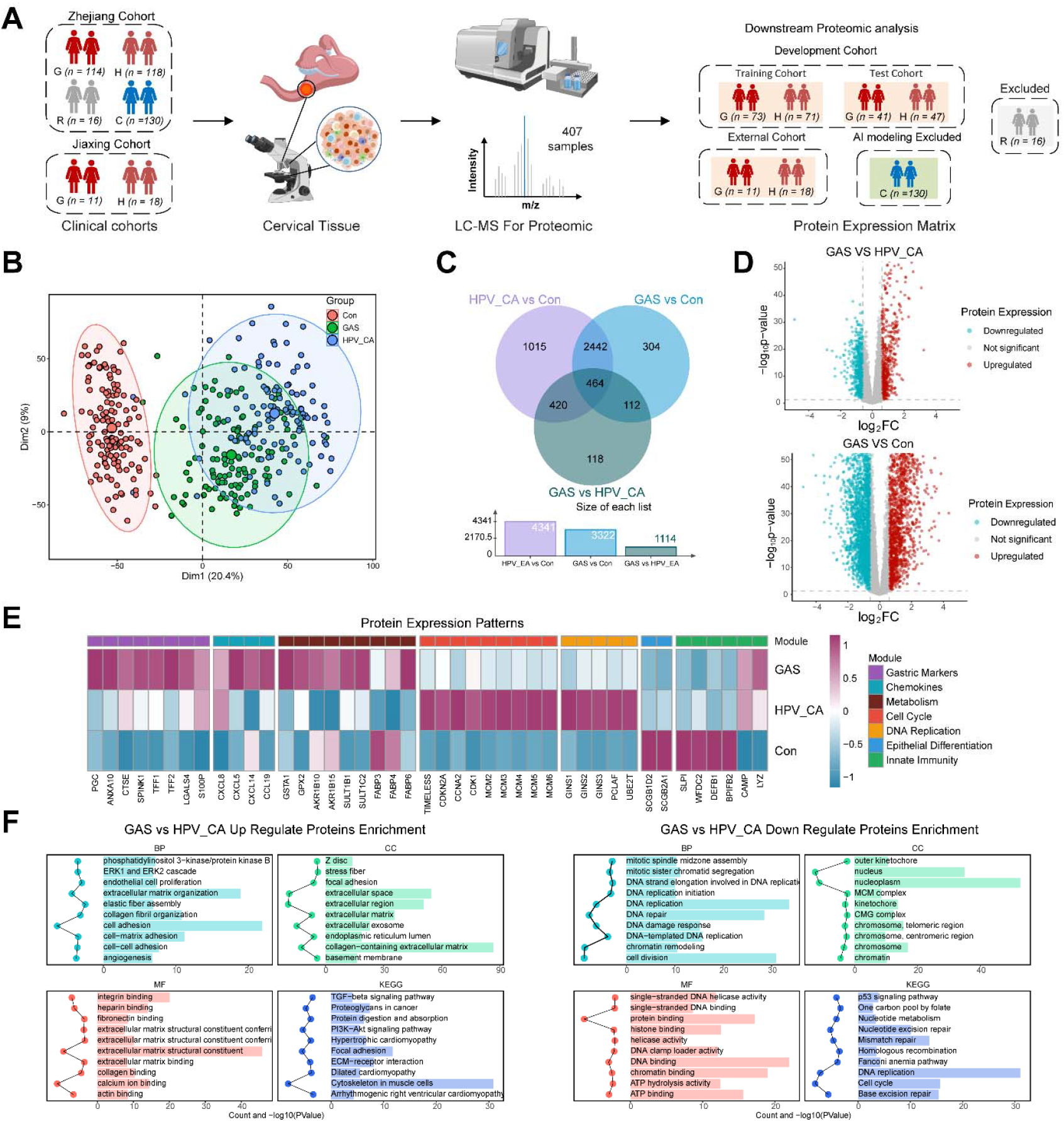
Comprehensive proteomic profiling reveals distinct molecular signatures across gastric adenocarcinoma subtypes. (**A**) Overview of study design and workflow. A multi-center cohort of 407 participants was recruited, comprising GAS (n = 125), HPV-CA (n = 136), rare non-GAS subtypes (n = 16), and control samples (n = 130). All 407 samples underwent LC-MS proteomic analysis. Subsequently, the rare subtypes (n = 16) and control samples (n = 130) were excluded from the deep learning diagnostic model construction. The remaining 261 disease samples were stratified into an independent External Proteomic Cohort (n = 29) and a Development Cohort (n = 232), with the latter being further subdivided into Training (n = 144) and Test sets (n = 88). (**B**) Principal component analysis (PCA) of proteomic profiles from control, GAS, and HPV-CA groups. (**C**) Venn diagram showing overlapping differentially expressed proteins among three pairwise comparisons: HPV-CA vs Con, GAS vs Con, and GAS vs HPV-CA. (**D**) Volcano plots displaying differentially expressed proteins for GAS vs HPV-CA (upper panel) and GAS vs Con (lower panel). Red dots represent downregulated proteins, cyan dots represent upregulated proteins, and gray dots represent non-significant proteins. (**E**). Heatmap showing protein expression patterns across sample groups (GAS, HPV-CA, and Con) for different modules. (**F**). Gene enrichment analysis for GAS vs HPV-CA comparison. Left panel shows enriched biological processes (BP), cellular components (CC), molecular functions (MF), and KEGG pathways for upregulated proteins. Right panel shows enrichment for downregulated proteins.

High-depth proteomic profiling achieved extensive coverage, quantifying an average of 9,036 proteins (8,992 protein groups) per sample, with protein identification and quantification filtered at a 1% false discovery rate (FDR) (Data S1). To ensure multi-batch dataset robustness, we executed a batch design using 8 continuous technical blocks integrated with 16 instrument pool QCs and 8 method pool QCs (fig. S1A). Cross-batch Pearson correlation analysis demonstrated exceptional reproducibility, with coefficients consistently exceeding 0.94 for both instrument and process controls, confirming a data structure with minimal batch effects (fig. S1B, C).

Principal component analysis (PCA) revealed that while malignant tissues separated cleanly from normal controls, the global expression profiles of GAS and HPV-CA exhibited extensive spatial overlap (Fig. 1B). Stratified PCA distributions across individual data splits, separate institutional cohorts, and combined clinical-origin groups consistently verified this morphological mimicry and shared malignant matrix (fig. S2A-F), indicating that traditional linear dimensionality reduction cannot decode the biochemical variances between these two adenocarcinoma subtypes.

We performed differential expression protein (DEP) analysis using an empirical Bayes moderated t-test (|Fold change| > 1.5 and Benjamini-Hochberg FDR-adjusted p-value < 0.05). We identified 4,341, 3,322, and 1,114 differentially expressed proteins (DEPs) in comparisons of HPV-CA vs. Con, GAS vs. Con, and GAS vs. HPV-CA, respectively (Fig. 1C, D). A cross-cohort overlay confirmed that these global landscape signatures remained stable across individual subsets (fig. S2G, H). Compared to the profound proteomic shifts against normal baselines, the restricted number of DEPs between GAS and HPV-CA highlights their interwoven oncogenic background and shared cellular derivation as cervical-derived tumors. Crucially, we used only training cohort-level DEPs solely for descriptive biological functional characterization, baseline pathway enrichment, and module-topology mapping. To ensure methodological rigor and prevent information leakage, we nested feature extraction, signature ranking, and hyperparameter tuning for the downstream WEDGE diagnostic modeling strictly and independently within the Training Cohort alone.

Functional module analysis further delineated the specific protein reprogramming in GAS across key biological dimensions (Fig. 1E). GAS expressed gastric differentiation markers, including PGC, CTSE, ANXA10, and TFF1/2, indicating that these cells retain the specialized secretory functions of gastric glandular epithelium through an HPV-independent differentiation mechanism. The immune microenvironment of GAS exhibited a robust inflammatory signature featuring chemokines such as CXCL8, CXCL5, CXCL14, and CCL19, reflecting extensive immune cell infiltration. We observed marked upregulation of stress-response and detoxification enzymes, particularly GSTA1, GPX2, and members of the AKR1B family (AKR1B10/15), in metabolic reprogramming in GAS.

Subsequent pathway enrichment analysis (GO and KEGG) provided mechanistic context to these expression patterns (Fig. 1F). Aligning with its high invasiveness, GAS showed significant upregulation of extracellular matrix remodeling, angiogenesis, and TGF-β signaling pathways, providing a molecular basis for its metastatic potential and poor prognosis. Furthermore, the enrichment of “protein digestion and absorption” pathways corroborated the gastric-type molecular signature identified in the single-protein analysis. Crucially, while HPV-CA was driven by DNA replication and cell-cycle pathways, GAS progression depended more on stromal remodeling and metabolic adaptation. Together, these results demonstrate that GAS exhibits profound proteomic reprogramming that reflects distinct differentiation states and biological processes and distinguishes it from HPV-related cervical adenocarcinoma.

### WEDGE framework for robust biomarker discovery

To overcome the high-dimensional noise and data sparsity inherent to rare-disease proteomics, we developed a computational framework termed WGAN-GP Enhanced Dual-stream GCN for Expression-Matrix (WEDGE) for classification and biological interpretability in rare-cancer proteomic matrices. The WEDGE framework integrates two synergized algorithmic modules: a Wasserstein Generative Adversarial Network with Gradient Penalty (WGAN-GP) for non-linear data augmentation, and a biologically informed Dual-stream Graph Convolutional Network (D-GCN) for deep feature extraction and network-based disease classification (Fig. 2A).

**Fig. 2.**
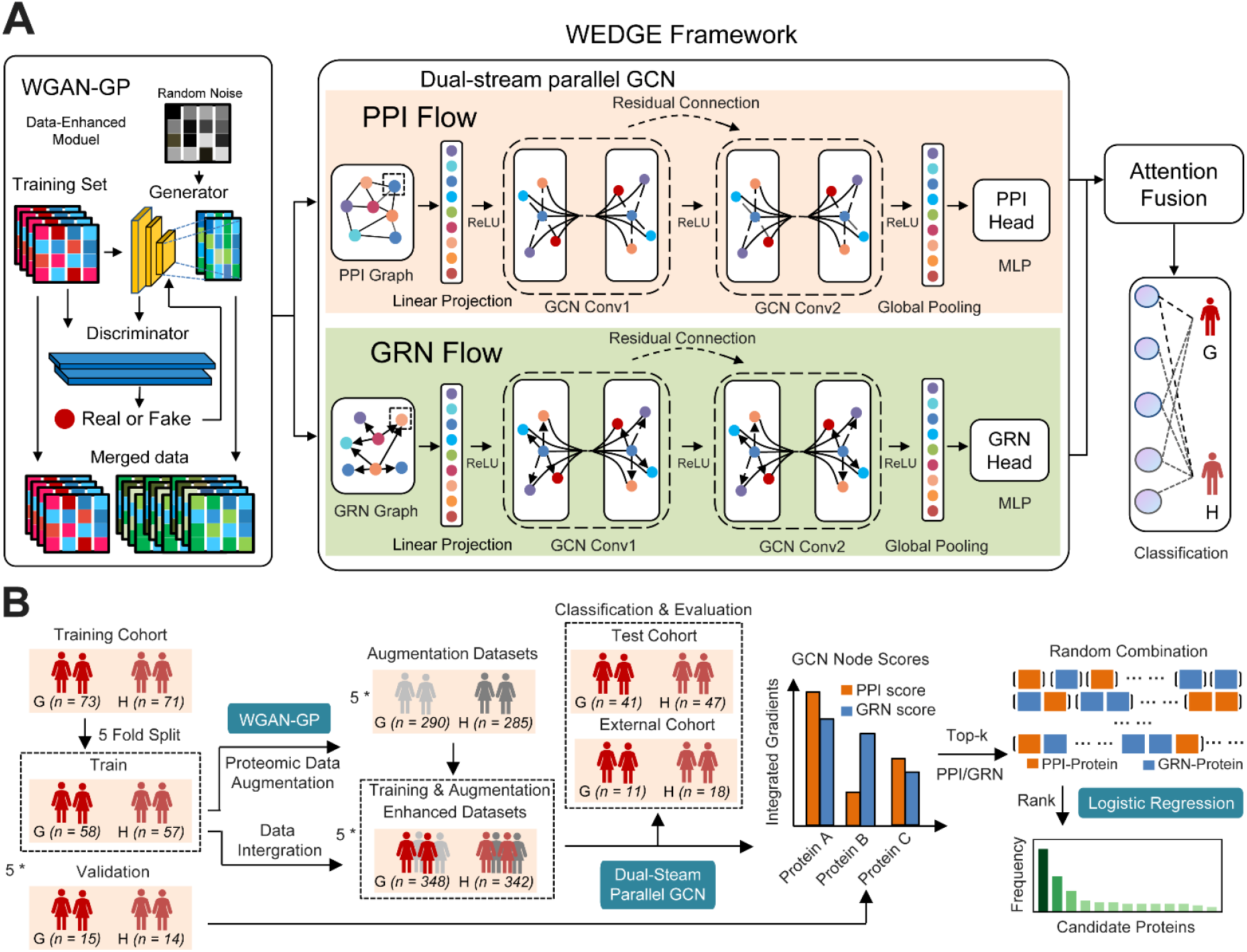
The overview of the WEDGE framework for GAS biomarker discovery. (**A**) The framework integrates WGAN-GP data augmentation with a D-GCN network. WGAN-GP generates enhanced datasets from real protein expression data. The D-GCN processes biological PPI and GRN networks simultaneously through parallel flows, where patient samples are represented as graphs with proteins as nodes, and the outputs converge into a classification module for disease prediction. (**B**) The workflow uses training cohorts (n = 144) to generate augmentation datasets. Enhanced datasets (n = 690) from WGAN-GP are utilized to train the D-GCN and are tested in Test Cohort and External Cohort. We then performed protein importance analysis and logistic regression to identify optimal biomarker combinations for GAS diagnosis. (G: GAS Patient; H: HPV-CA Patient)

In the WGAN-GP module, a generator and a discriminator are trained on real protein expression matrices, where the discriminator guides the generator to produce high-quality synthetic data. The resulting synthetic samples are then merged with the real dataset to feed the downstream D-GCN network. D-GCN embeds topological priors into the deep learning process, transforming flat protein matrices into graph-structured functional networks in which each protein serves as a node (Fig. 2A). Specifically, before graph propagation, stream-specific linear projection layers map each protein’s scalar expression into a continuous embedding. This projection expands feature expressiveness and aligns the embeddings with the downstream streams. To capture complementary dimensions of tumor biology, the framework processes two parallel networks: the PPI Flow aggregates undirected physical interactome coordinates and co-functional protein-protein interactions, while the GRN Flow captures the directed, unidirectional gene regulatory networks (Fig. 2A). Within each stream, graph convolutional layers with residual connections propagate node features across these biological topologies. A learnable attention fusion mechanism integrates the intermediate representations from both streams, dynamically weighting their contributions to predict disease categories (Fig. 2A).

To implement this architecture, we followed a multi-stage nested workflow (Fig. 2B). We optimized the framework using stratified 5-fold cross-validation restricted to the Training Cohort (n = 144; 73 GAS and 71 HPV-CA), keeping the Internal Test Cohort (n = 88) and External Proteomic Cohort (n = 29) entirely untouched throughout the pipeline (Fig. 2B). To prevent data leakage, we executed the WGAN-GP augmentation strictly within the inner loop of each cross-validation fold: for each iteration, the module trained exclusively on the 4/5 training partition to generate five synthetic instances per real sample (yielding 575 simulated profiles per fold), while we held the remaining 1/5 validation fold unaltered for unbiased evaluation (Fig. 2B). We scaled the synthetic profiles to match the original proteomic scale and merged them with the authentic samples, yielding 690 instances per fold (348 GAS and 342 HPV-CA) to stabilize training (Fig. 2B).

To distill the high-dimensional graph representations into diagnostic features, we used a three-step feature attribution and consensus selection protocol (Fig. 2B). First, we applied Integrated Gradients to the optimized GCN weights to extract the top 15 highest-scoring features from each flow, yielding 30 core network-interactive candidate features. Second, to identify the most parsimonious predictor panel, we evaluated combinations of 3-10 proteins from the 30-protein pool using Logistic Regression within the nested 5-fold cross-validation on the training cohort. Tracking panel size against performance variance showed that a two-protein combination formed the optimal baseline, with larger panels providing no additional statistical benefit. Third, we ranked combinations solely by their validation-fold metrics and tracked the recurrence frequency of each protein across the top-performing ensembles (Fig. 2B). This frequency-based selection filtered out single-feature dominance and co-selection noise, yielding a stable dual-biomarker panel.

### WGAN-GP augmentation validation

To establish the stability of the computational framework in small-cohort settings, we evaluated the optimization dynamics and distributional fidelity of the WGAN-GP module. We monitored the training trajectories for GRN, PPI, and Total Loss across all five cross-validation loops and confirmed stable neural network optimization without structural variance (fig. S3A-C).

To quantify the fidelity of the synthesized proteomic profiles, we evaluated their Fréchet Distance (FD) against real patient matrices. The WGAN-GP-generated profiles achieved low FD scores across all loops (471.05-547.33), outperforming the randomized Gaussian noise baseline (1201.08) and confirming that the generative pipeline captured genuine high-dimensional molecular dependencies rather than background noise (Fig. 3A). Across folds, the Fold 5 configuration achieved the lowest validation loss (0.3654) and the lowest FD (471.05), identifying it as the optimal architecture for preserving authentic proteomic structure. While classification accuracy varied slightly across folds (Fig. 3B), the dual minimization of validation loss and FD led us to select Fold 5 as the discovery backbone for generating stabilized training matrices and serving as the unbiased template for downstream Integrated Gradients attribution. PCA showed that the augmented samples co-localized with authentic samples in the first two principal components while preserving the inter-subtype separation (Fig. 3C). The Fold 5 WGAN-GP-generated synthetic profiles are deposited in Data S2.

**Fig. 3.**
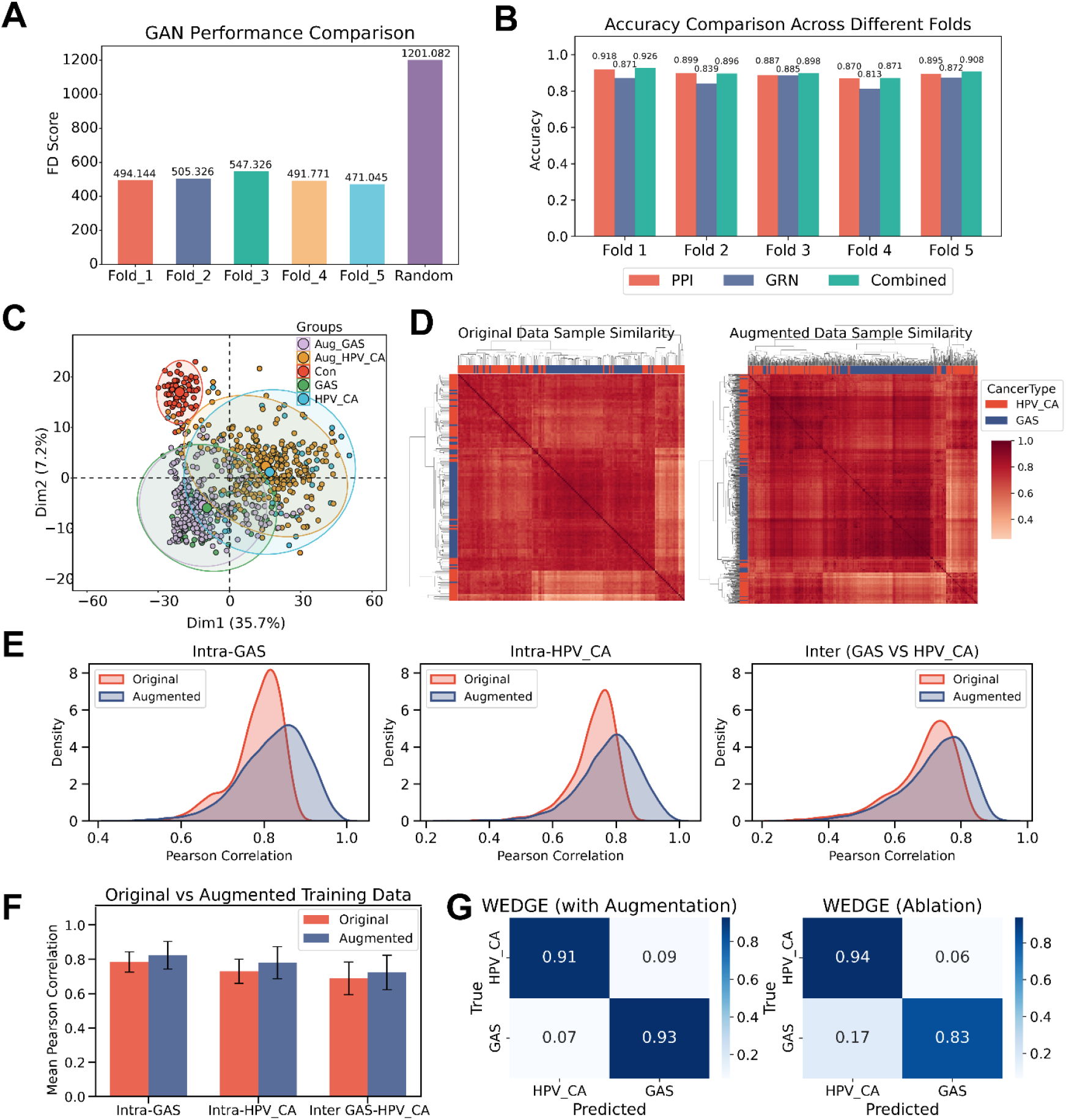
Evaluation of WGAN-GP data augmentation. (**A**) Fréchet Distance (FD) scores comparing GAN-generated data quality across five folds versus random baseline. (**B**) Accuracy comparison across different folds using PPI network, GRN network, and combined dual-stream approach. (**C**) Principal component analysis (PCA) visualization of augmented GAS, augmented HPV-CA, original control, original GAS, and original HPV-CA samples. (**D**) Hierarchical clustering heatmaps representing sample similarity (Pearson correlation) within the original data (left) and the augmented data (right). (**E**) Probability density plots (KDE) of Pearson correlation coefficients for Intra GAS, Intra HPV-CA, and Inter-group (GAS vs. HPV-CA) comparisons between original and augmented datasets. (**F**) Bar chart displaying the mean Pearson correlation coefficients of original versus augmented training data across intra and inter-group relations. (**G**) Confusion matrices demonstrating the classification performance of the WEDGE model (with data augmentation, left) versus the ablation model (without data augmentation, right) in separating HPV-CA and GAS groups. Note that the classification is based on end-to-end patient graph embeddings rather than downstream selected protein biomarkers.

We further validated the fidelity of the synthetic profiles through sample-to-sample similarity and correlation topology analysis. Hierarchical clustering showed that the generated profiles closely mirrored the real samples in global structure (Fig. 3D). Probability density distributions and mean correlation trajectories further revealed that WGAN-GP augmentation addressed the limited resolution of the authentic dataset (Fig. 3E, F). In the original data, the gap (step-gradients) between intra-class and inter-class mean Pearson r was 0.095 for GAS and 0.041 for HPV-CA, highlighting the overlap that complicates classification. Following inner-loop augmentation, these boundaries widened to 0.101 and 0.056, respectively (Fig. 3F). This expansion showed that generative enhancement broadened class discriminability without introducing spurious overlap.

A full-scale classification ablation underscored the necessity of generative data augmentation: the full WEDGE model achieved recall of 0.91 for HPV-CA and 0.93 for GAS (Fig. 3G). Omitting the WGAN-GP enrichment module (WEDGE Ablation) reduced performance specifically for the rare lineage, with GAS recall dropping to 0.83 because of minority-class under-representation, confirming that inner-loop data augmentation eliminates classification bias (Fig. 3G).

### Feature selection and optimal biomarker identification

To decode the structural topologies driving the dual-stream network decisions, we evaluated node feature attributions using an inner-loop validation framework via Integrated Gradients, mapping importance weights back to specific protein coordinates. We systematically evaluated feature importance within the trained D-GCN to identify key discriminatory proteins.

In the PPI network, PGC emerged as the top-ranked feature (score: 10.00), complemented by extracellular and glandular differentiation modulators including SBSN (7.74), CTSE (6.85), and POTEKP (6.14) (Fig. 4A). Concurrently, the directed control cascade (GRN Flow) prioritized upstream transcriptional regulators, ranking ACTA2 (10.00), FOXA1 (7.80), and BRCA1 (5.69) as leading functional contributors alongside the clinical epigenetic checkpoint DNMT1 (5.06) and ELF3 (4.49) (Fig. 4A). Uniting the top 15 highest-scoring features from both streams formed a highly condensed candidate pool of 30 network-interactive nodes (Data S3). Notably, CDX2 (3.21) is consistent with prior IHC-based reports designating it as a potential biomarker for GAS detection(*27*). To benchmark the diagnostic efficacy of WEDGE against established biomarker discovery paradigms, we performed comparison with four frameworks: BINN(*28*)(deep learning), POC-19(*29*) (machine learning panel), RF (Random Forest), and DIABLO (bioinformatics, latent component integration) (*30*). Evaluation across varying signature sizes (1 to 15 proteins) revealed that WEDGE achieved superior classification accuracy compared to all baseline methods (Fig. 4B). Notably, at an ultra-compact dimension of just two proteins, WEDGE attained a dominant classification accuracy of 0.93, substantially outperforming BINN (0.83), POC-19 (0.82), DIABLO (0.89), and RF (0.90), establishing the high feature-extraction efficiency of our graph-regularized learning architecture.

**Fig. 4.**
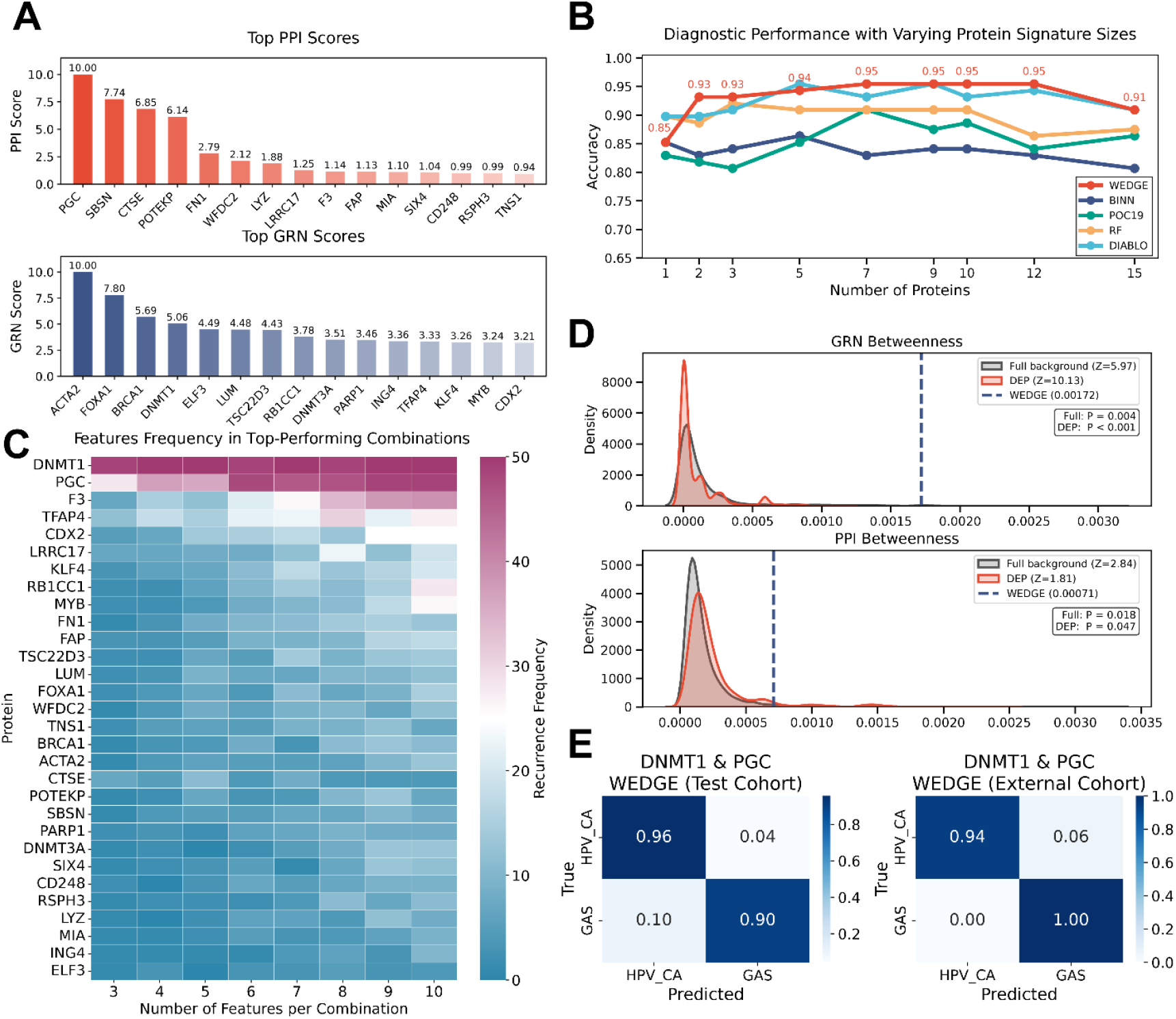
Feature selection, identification of the optimal biomarker panel, and network topological validation. (**A**) Bar charts displaying the ranking of candidate features based on Top PPI Scores (top) and Top GRN Scores (bottom) to identify highly connected network nodes. (**B**) Line graph comparing the diagnostic accuracy of WEDGE against other benchmark methods (BINN, POC19, RF, DIABLO) across varying numbers of protein signatures (from 1 to 15). (**C**) Heatmap illustrating the recurrence frequency of specific proteins within the top-performing feature combinations across different signature sizes (from 3 to 10 features per combination), highlighting DNMT1 and PGC as the most stable features. (**D**) Permutation test density plots (KDE) of the null distributions for GRN Betweenness (top) and PPI Betweenness (bottom). The dashed vertical lines represent the observed means of the WEDGE panel against the full universe (grey) and the DEP set (orange) backgrounds. (**E**) Confusion matrices for the optimal two-protein biomarker panel (DNMT1 & PGC) evaluated by WEDGE, demonstrating robust classification performance in both the test cohort (left) and the independent External Proteomic Cohort (right).

To identify robust constituents for the signature, we used consensus selection: we evaluated random combinations from the 30-protein pool, ranked them by validation-fold metrics, and tracked the recurrence frequency of each protein across the top 50 models per panel size (Fig. 4C). This analysis showed that DNMT1 and PGC dominated: they were the most frequently selected proteins across panel sizes of 3-10 (Fig. 4C). Evaluating panel size against validation variance, we finalized a two-protein panel (PGC and DNMT1) as the parsimonious baseline, with larger panels providing no additional benefit.

To assess mechanistic plausibility, we evaluated the topological properties of PGC and DNMT1 within PPI and GRN networks via 10,000-iteration permutation tests (Fig. 4D). WEDGE-identified biomarkers showed high network centrality compared with both the full network and the DEP background. Specifically, the observed mean GRN Betweenness of the panel was enriched against both background sets (Full background: Empirical p-value = 0.004, Z=5.97; DEP background: Empirical P < 0.001, Z=10.13; Fig. 4D). Additionally, the panel’s PPI Betweenness demonstrated a modest but significant topological enrichment (Full background: Empirical p-value = 0.018, Z=2.84; DEP background: Empirical p-value = 0.047, Z=1.81; Fig. 4D). This analysis shows that PGC and DNMT1 are key network hubs, supporting their diagnostic relevance.

To confirm the biological plausibility of the PGC-DNMT1 panel, we performed expression profiling across the clinical cohorts. Unsupervised PCA using only these two markers separated GAS and HPV-CA into distinct clusters, reproducing the separation pattern seen across the full proteome (fig. S4A). The SHAP summary plot for the WEDGE model showed the relative contributions and directions of PGC and DNMT1 on the model output (fig. S4B). Quantitative mass-spectrometry-derived expression profiles confirmed that PGC was upregulated in GAS while DNMT1 was elevated in HPV-CA (***p < 0.001) (fig. S4 C-D). Latent-distribution matching showed that WGAN-GP-generated profiles for PGC and DNMT1 closely matched the empirical distributions of the real patient cohorts in both HPV-CA and GAS classes (fig. S4 E-F). Single-marker classification with PGC alone achieved 94%/76% recall for HPV-CA/GAS, while DNMT1 alone achieved 89%/80%. Both single markers showed respectable but imbalanced discriminatory capacity compared with the balanced two-protein panel (fig. S4 G-H). The combined PGC-DNMT1 panel performed strongly across both validation cohorts. In the Internal Test Cohort (n = 88), WEDGE achieved recall of 0.96 for HPV-CA and 0.90 for GAS, and in the External Proteomic Cohort, recall of 0.94 for HPV-CA and 1.00 for GAS (Fig. 4E). These dual-cohort results show that the WEDGE discovery framework generalizes across centers for the differential diagnosis of cervical adenocarcinoma subtypes.

### WEDGE outperforms existing biomarker discovery methods for GAS classification

To benchmark the diagnostic efficacy of the WEDGE-derived signature, we compared its performance against four established algorithms (BINN, POC-19, RF, and DIABLO). We restricted all models to their top-two ranked proteins to enable fair comparison of feature selection efficiency.

In the Internal Test Cohort, WEDGE achieved the highest overall accuracy (0.93) and an AUC of 0.98, outperforming DIABLO (0.89), RF (0.90), POC-19 (0.82), and BINN (0.83) (Fig. 5A, B). Stratified metric profiling highlighted WEDGE’s balanced diagnostic capacity, achieving coordinated recall and precision rates across both the dominant HPV-CA lineage and the rare, morphologically mimetic GAS lineage (Fig. 5C, D). By contrast, POC-19 and BINN showed markedly lower GAS recall under the limited training data (Fig. 5E).

**Fig. 5.**
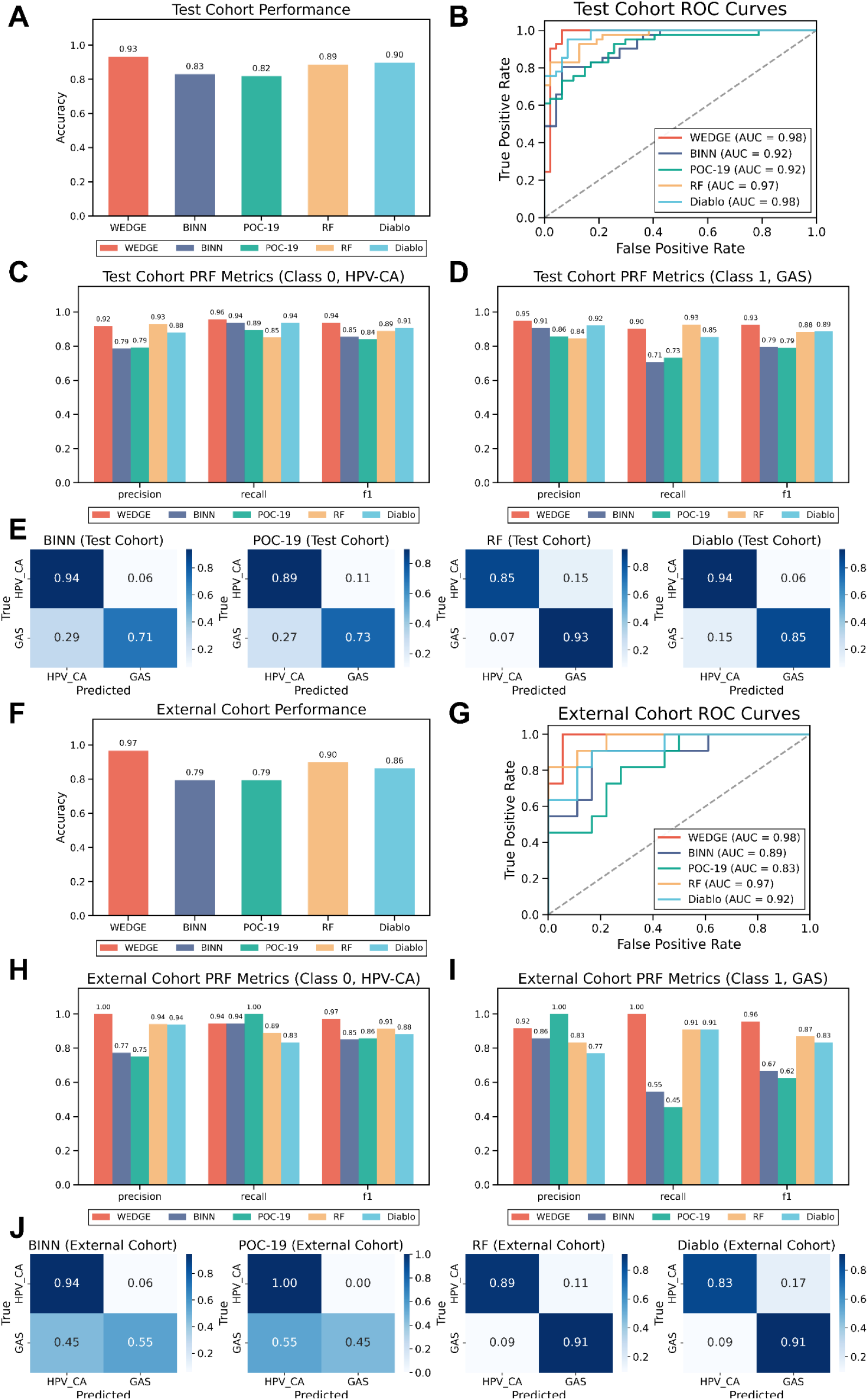
Performance comparison of WEDGE with alternative biomarker discovery methods. (**A**) Bar chart comparing the accuracy of WEDGE against BINN, POC-19, RF, and Diablo in the Internal Test Cohort. (**B**) Receiver Operating Characteristic (ROC) curves displaying the Area Under the Curve (AUC) for all methods in the Test Cohort. (**C**) Precision, Recall, and F1-score metrics for Class 0 (HPV-CA) in the Test Cohort. (**D**) Precision, Recall, and F1-score metrics for Class 1 (GAS) in the Test Cohort. (**E**) Confusion matrices showing the classification performance of comparative methods (BINN, POC-19, RF, and Diablo) in the Internal Test Cohort. (**F**) Bar chart comparing the accuracy of WEDGE against other methods in the External Cohort. (**G**) ROC curves displaying the AUC for all methods in the External Cohort. (**H**) Precision, Recall, and F1-score metrics for Class 0 (HPV-CA) in the External Cohort. (**I**) Precision, Recall, and F1-score metrics for Class 1 (GAS) in the External Cohort. (**J**) Confusion matrices showing the classification performance of comparative methods (BINN, POC-19, RF, and Diablo) in the External Cohort.

In the cross-center External Proteomic Cohort, WEDGE achieved an overall accuracy of 0.97 and an AUC of 0.98, outperforming DIABLO (0.93), RF (0.83), and both BINN and POC-19 (0.79 each) (Fig. 5F, G). Class-specific evaluation showed that WEDGE reached a precision of 1.00 for HPV-CA (Fig. 5H) and a recall of 1.00 for the rare GAS lineage (Fig. 5I). By contrast, the conventional classifiers showed substantial performance drops across centers (Fig. 5J): POC-19 collapsed into a single-class predictor biased toward HPV-CA (recall: 1.00 for HPV-CA, 0.45 for GAS), and BINN showed a similar pattern (recall: 0.55 for GAS) (Fig. 5J). Together, these results indicate that WEDGE provides more stable classification than conventional classifiers in small-cohort, multi-center settings.

### Immunohistochemistry (IHC) validation of WEDGE-identified biomarkers

To corroborate the clinical translatability of the WEDGE-identified biomarkers, we performed immunohistochemical (IHC) validation in an independent IHC validation cohort encompassing formalin-fixed paraffin-embedded (FFPE) tissue sections, comprising 50 GAS, 60 HPV-CA, and 15 morphologically normal control samples. This cohort represents a distinct patient population that does not overlap with the proteomic discovery or external validation cohorts.

To bridge the methodological gap between the continuous scalar matrix of mass spectrometry and the semi-quantitative, discrete ordinal scale of clinical pathology, we deployed an XGBoost (eXtreme Gradient Boosting)-based machine learning architecture trained on pathologist-mediated IHC staining scores (0-2 intensity scale) (Fig. 6A). For robust model development and unbiased evaluation, the 110 adenocarcinoma specimens were partitioned into an IHC training set (n = 77; 35 GAS and 42 HPV-CA) and IHC test set (n = 33; 15 GAS and 18 HPV-CA), while the 15 normal control samples were utilized exclusively to establish baseline physiological expression profiles (Fig. 6A). A semi-quantitative staining scoring system was established for the two-protein panel (PGC and DNMT1), evaluating both staining intensity and extent in each patient sample (Data S4).

**Fig. 6.**
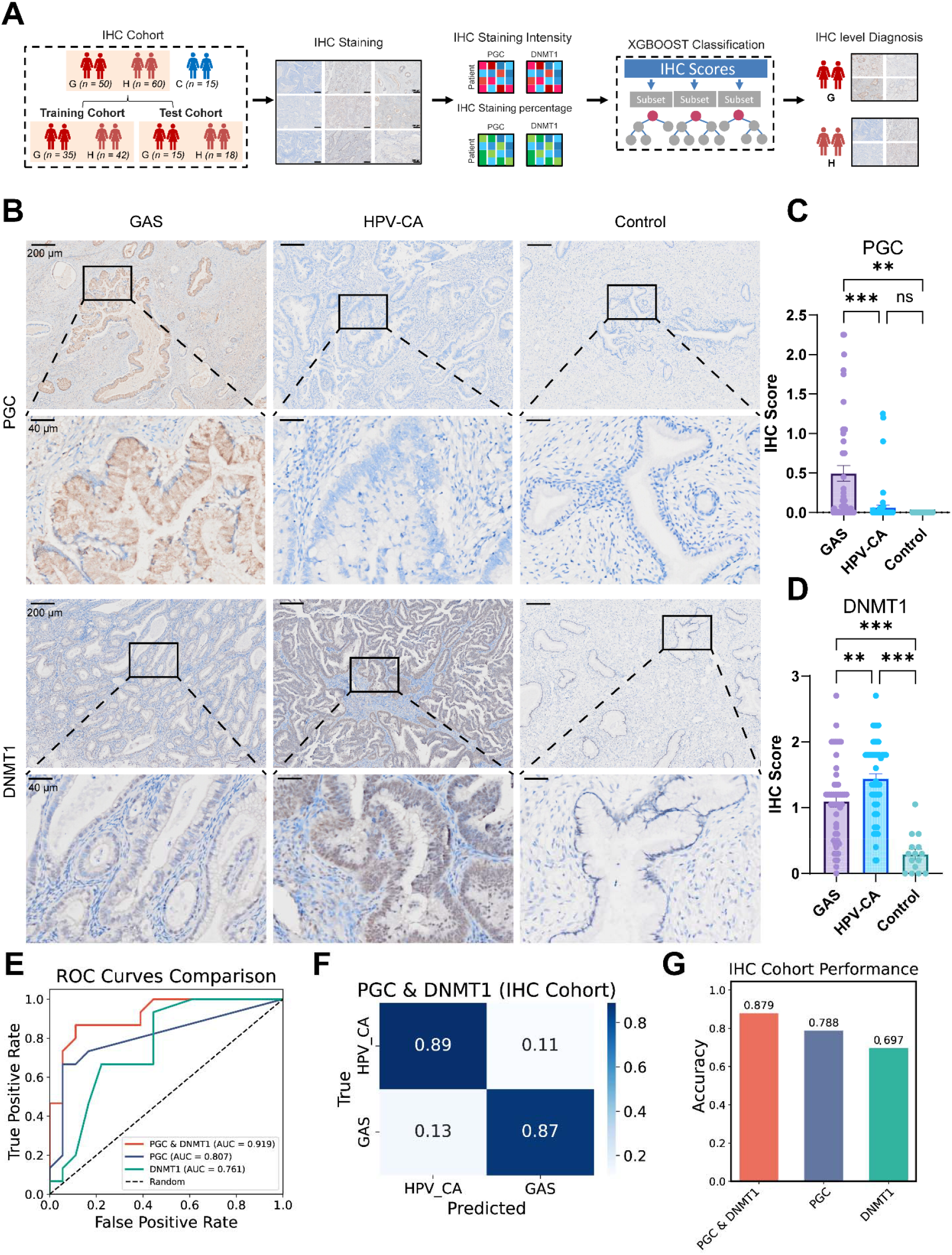
IHC validation of WEDGE-identified biomarkers for GAS and HPV-CA classification. (**A**) Workflow of the IHC validation and diagnostic model construction. An XGBoost machine learning classifier was trained using IHC scores derived from tissue samples of 50 GAS cases and 60 HPV-CA cases, with 15 control samples included for expression baseline analysis. (**B**) Quantitative comparison of IHC scores for the two biomarkers (PGC and DNMT1) across GAS, HPV-CA, and Control groups. (**C**) Comparison of IHC scores for PGC. Statistical significance was determined using Tukey’s multiple comparisons test. (***p < 0.001, **p < 0.01, *p < 0.05, ns: not significant) (**D**) Comparison of IHC scores for DNMT1. Statistical significance was determined using Tukey’s multiple comparisons test. (***p < 0.001, **p < 0.01, *p < 0.05, ns: not significant). (**E**) ROC curves comparing the diagnostic performance of different biomarker combinations in the IHC cohort. The enlarged single-marker ROC curves for PGC alone and DNMT1 alone are shown in fig. S5F, G to enable clear inspection of their respective AUC values. (**F**) Confusion matrix showing the classification performance of the two-protein panel model. (**G**) Bar plot comparing the diagnostic accuracy of different biomarker combinations.

Morphological evaluation revealed distinct staining patterns for the identified markers. In GAS tissues, PGC demonstrated medium-intensity cytoplasmic staining within the tumor cells, typically observed in regions of prominent stromal sclerosis. Two senior gynecological pathologists independently scored the staining, with disagreements resolved by consensus. In contrast, DNMT1 in HPV-CA samples showed distinct nuclear localization, with tumor cell nuclei displaying medium-intensity staining against the blue-stained tumor stroma, providing clear internal contrast (Fig. 6B). IHC scoring confirmed significant differential expression across the groups. PGC showed a highly discriminatory profile with near-exclusive positivity in GAS patients compared to both HPV-CA and control groups (Fig. 6C). Although DNMT1 expression was present in both adenocarcinoma subtypes, its staining intensity was significantly higher in HPV-CA than in GAS and control samples (Fig. 6D). These differential patterns were further supported by visual inspection of representative staining images and comprehensive intensity heatmaps (fig. S5A). In the diagnostic validation, the two-protein panel (PGC & DNMT1) demonstrated robust performance, achieving an AUC of 0.919 (Fig. 6E). Comprehensive performance metrics further confirmed that the combined panel provided superior and more consistent results across both diagnostic categories compared to single-protein models (fig. S5B, C). The corresponding classification performance visualised in the confusion matrices (fig. S5D, E). ROC analysis confirmed that the combined PGC-DNMT1 panel achieved superior discriminative capacity over either single marker (PGC: AUC = 0.807; DNMT1: AUC = 0.761; single-marker curves enlarged in fig. S5F, G) (Fig. 6E). The integrated XGBoost model correctly identified 87% of GAS cases and 89% of HPV-CA cases (Fig. 6F), resulting in an overall accuracy of 0.879 (Fig. 6G). Together, these analyses confirmed that the combination of PGC and DNMT1 effectively harnessed their complementary expression profiles to optimize diagnostic accuracy. These multi-center clinical validation outcomes definitively confirm that the WEDGE-derived biomarker panel can be seamlessly integrated into standard diagnostic pathology workflows, providing an accessible and highly reliable solution for resolving these morphologically mimetic cervical malignancies.

### Construction and evaluation of a PGC-based prognostic risk model

As diagnostic biomarkers often reflect the underlying biological aggressiveness and therapeutic resistance of tumors, they frequently also harbor latent prognostic significance(*31*). Given that PGC was identified as a top-ranked feature in the WEDGE framework and demonstrated the high diagnostic specificity in the IHC validation, we next investigated its potential prognostic significance. The prognostic evaluation was restricted to PGC, rather than DNMT1, because DNMT1 is primarily upregulated in HPV-CA (where it reflects virus-driven epigenetic remodeling), whereas the survival analysis was confined to the GAS cohort, in which PGC is the dominant lineage-specific marker with biologically meaningful variation. Kaplan-Meier analysis on 95 GAS patients with complete follow-up information revealed that PGC expression was significantly associated with overall survival (OS), with PGC-positive patients exhibiting superior outcomes compared to those with negative expression (Fig. 7A). This was further supported by survival duration analysis, where the PGC-positive group showed significantly longer mean OS times (Fig. 7B). Parallel survival distributions for alternative standard clinicopathological parameters, including FIGO stage and parametrial involvement, are provided for baseline reference (fig. S6A, B).

**Fig. 7.**
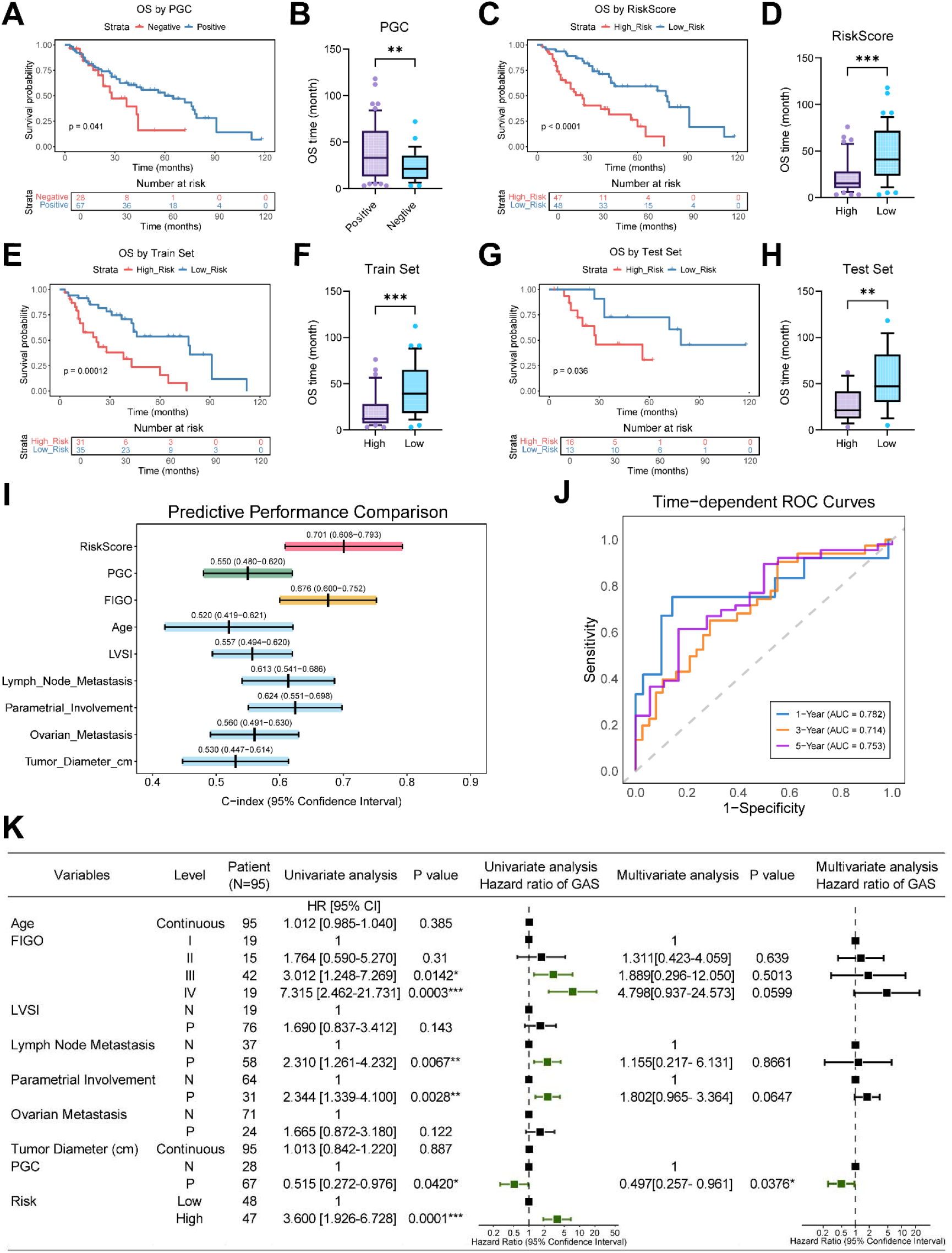
PGC biomarker for prognostic model. (**A**) Kaplan-Meier survival analysis of Overall Survival (OS) based on PGC expression status (Negative vs. Positive, n = 95). (**B**) Box plot showing the distribution of OS time (months) comparing PGC-positive and PGC-negative groups. (** p < 0.01). (**C**) Kaplan-Meier OS analysis stratified by the integrated Risk Score (Low Risk Group vs. High Risk Group, n = 95). (**D**) Box plot showing the distribution of OS time (months) comparing High-Risk and Low-Risk groups. (*** p < 0.001). (**E**) Kaplan-Meier OS analysis stratified by Risk Score in the Training Set. (**F**) Box plot showing the distribution of OS time (months) in the Training Set (High vs. Low Risk). (*** p < 0.001). (**G**) Kaplan-Meier OS analysis stratified by Risk Score in the Test Set. (**H**) Box plot showing the distribution of OS time (months) in the Test Set (High vs. Low Risk). (** p < 0.01). (**I**) Forest plot of Concordance Indices (C-index) comparing the predictive accuracy of the Risk Score against individual clinicopathological variables and PGC. Data are presented as C-index values with 95% confidence intervals. (**J**) Time-dependent ROC curve analyses of the prediction model for 1-, 3-, and 5-year survival outcomes. (**K**) Univariate and Multivariate Cox proportional hazards regression analyses identifying independent prognostic factors (n = 95). The table lists detailed statistics, and the forest plots display Hazard Ratios (HR) with 95% Confidence Intervals. (N = Negative, P = Positive; LVSI=Lymphovascular Space Invasion).

To translate this biological finding into a clinically applicable predictive tool, we further constructed a comprehensive risk model by integrating PGC expression with established clinicopathological variables. The resulting Risk Score effectively stratified the 95 patients into distinct risk groups, with the high-risk group showing significantly inferior OS (Fig. 7C) and significantly shorter survival durations (Fig. 7D). Consistent with this, the high-risk group was associated with a significantly higher mortality rate and shorter overall survival duration (p = 0.0076, Fig. 7D). The robustness of this risk model was validated across split cohorts, demonstrating consistent prognostic significance in both Training Set (Fig. 7E, F) and Test Set (p = 0.036, Fig. 7G, H).

We next evaluated the predictive precision of the model using the Concordance Index (C-index). The integrated Risk Score demonstrated the highest predictive accuracy, achieving a C-index of 0.701 (95% confidence interval [CI]: 0.608-0.793), outperforming FIGO stage (0.676), PGC status alone (0.550), and other individual clinicopathological variables (Fig. 7I). Time-dependent ROC analysis further substantiated the model’s reliability, yielding AUC values of 0.782 at 1-year, 0.714 at 3-year, and 0.753 at 5-year follow-up (Fig. 7J). Furthermore, a comprehensive temporal comparison against individual clinical factors (fig. S6C) revealed that while FIGO stage exhibited competitive prognostic value at the 3-year interval, the integrated Risk Score maintained superior predictive performance for both short-term (1-year) and long-term (5-year) survival outcomes, demonstrating its stability.

Finally, univariate and multivariate Cox regression analyses were performed to identify independent prognostic factors. In the univariate analysis, FIGO stage III/IV, lymph node metastasis (p = 0.0067), parametrial involvement (p = 0.0028), PGC expression (p = 0.042), and the Risk Score (p = 0.0001) were associated with OS (Fig. 7K). Variables with statistical significance in the univariate analysis were subsequently incorporated into a multivariate Cox proportional hazards model to adjust for potential confounding factors. Given that the Risk Score is a composite index integrating these clinicopathological features, it was deliberately excluded from the multivariate model to prevent multicollinearity. Crucially, multivariate analysis confirmed that PGC expression remained a robust independent prognostic factor (hazard ratio [HR] = 0.497, p = 0.0376). These findings suggest that PGC expression contributes meaningfully to patient survival outcomes. Alongside its diagnostic utility, PGC demonstrates potential as a prognostic indicator for GAS, providing valuable prognostic information that complements standard clinicopathological evaluations.

## DISCUSSION

GAS remains one of the most diagnostically challenging subtypes of cervical adenocarcinoma, owing to its morphological mimicry of benign endocervical glands and the limited sensitivity and specificity of currently available markers, issues that often translate into misclassification and delayed intervention(*5, 32*). To address this, we conducted a systematic multi-center proteomic analysis of GAS and developed the WEDGE framework for robust biomarker discovery. Through WEDGE and an IHC-based approach, we identified and validated a two-protein panel (PGC and DNMT1) with high diagnostic accuracy. Together with the prognostic value of PGC, these findings provide a clinically actionable framework for the diagnosis and management of this aggressive disease.

A central contribution of this work lies in the comprehensive proteomic delineation of GAS and its systematic comparison with HPV-CA. While previous genomic and transcriptomic studies have provided valuable insights (*33–35*), they were often constrained by limited cohort sizes and offered only indirect proxies of the functional tumor phenotype, hindering the discovery of robust diagnostic biomarkers. No comprehensive proteomic characterization of GAS has been reported to date, leaving a critical gap in understanding its functional phenotype and translational potential. Here, we leveraged a multi-center cohort to construct a high-resolution proteomic atlas that captures the functional state of GAS beyond genomic alterations. Our global proteomic characterization revealed a profound divergence in oncogenic drivers between GAS and HPV-CA. The proteomic landscape of GAS is defined by a specialized secretory reprogramming, characterized by robust upregulation of gastric-type modules including PGC, CTSE, and TFF1/2. This reflects an ectopic lineage commitment fundamentally distinct from the typical differentiation of the cervix. By contrast, the HPV-CA proteome is primarily fueled by hyper-proliferation machinery, showing significant enrichment in cell-cycle regulators and DNA replication components such as the MCM complexes. These findings indicate that HPV-CA is driven by constitutive proliferative signaling, while GAS progression is governed by a unique gastric-type differentiation program. Translating these expansive systems-level insights into robust, clinically actionable biomarkers remains a formidable task, as traditional statistical approaches are often insufficient to navigate the high-dimensional noise and inherent data sparsity of rare-disease cohorts. To bridge this gap between global proteomic characterization and precise diagnostic application, we developed the WEDGE framework to resolve the “curse of dimensionality” (p ≫ n) that has long constrained rare-disease research (*36, 37*). In small-cohort studies, the high-dimensional feature space often leads to “spurious correlations” where traditional statistical models capture noise rather than true biological signals (*38*). At the data layer, WGAN-GP is used for data augmentation. Unlike classical oversampling methods such as SMOTE(*39*), which typically interpolate in feature space and may fail to account for the complex, non-linear geometry of biological data, WGAN-GP generates new samples by learning the underlying latent distribution (*40, 41*). Our ablation studies further empirically demonstrated this necessity: omission of the WGAN-GP module directly led to a drop in GAS recall (from 0.93 to 0.83), thereby validating its power in neutralizing classification biases inherent to rare diseases. At the model layer, WEDGE embeds biological inductive bias into learning by representing proteins as nodes in protein-protein interaction (PPI) and gene-regulatory (GRN) networks within a dual-stream graph convolutional network. This design shifts the modeling paradigm from a flat vector of independent features to message passing across biological topologies (*42*). This allows the model to aggregate features based not only on individual abundance, but also on the functional context of each protein within the network. The PPI flow captures the stable machinery of the physical protein complexes and co-functional modules, while the GRN flow captures the dynamic regulatory cascades that drive malignant transformation (*43–46*). This multi-view learning strategy treats GAS as a systemic failure of cellular programs rather than a collection of isolated protein alterations.

By synthesizing these complementary perspectives through an attention mechanism, WEDGE identifies higher-order interactions that are largely invisible to univariate statistical tests, ultimately distilling the global proteomic variance into a streamlined two-protein diagnostic panel: PGC and DNMT1. The biological validity of this lineage-based selection is further reinforced by our network-based identification of CDX2, a previously reported IHC marker for GAS, which confirms the model’s ability to capture authentic gastric-differentiation signals (*27*). The molecular characterization of the PGC-DNMT1 panel underscores a fundamental divergence in oncogenic drivers linking lineage identity and epigenetic regulation. PGC, a canonical secretory marker of gastric chief cells, exhibits robust expression in GAS (*47, 48*), indicating it as a molecular footprint of the unique gastric lineage reprogramming inherent to this malignancy. By contrast, the differential expression of DNMT1 provides a critical surrogate for the HPV-driven status of these adenocarcinoma subtypes. Extensive evidence demonstrates that HR-HPV E6 and E7 oncoproteins constitutively upregulate DNMT1, either by preventing its proteasomal degradation or through p53-mediated pathways, to facilitate viral persistence and silence host tumor suppressors via aberrant DNA methylation (*49*). Consequently, the significantly higher DNMT1 levels in HPV-CA reflect a state of viral-mediated epigenetic remodeling, whereas the relatively lower expression in GAS highlights an HPV-independent oncogenic trajectory.

To bridge the gap between systems-level insights and clinical practice, our IHC validation in the IHC validation Cohort demonstrates that the proteomic signatures identified by WEDGE are translatable to routine diagnostic settings. Morphological evaluation confirmed that the molecular divergence of these entities manifests as distinct staining patterns, with PGC showing cytoplasmic expression in GAS and DNMT1 showing nuclear localization in HPV-CA, offering pathologists clear and contrasting visual cues for differential diagnosis. The integrated model significantly outperforms conventional single-biomarker assessments. Widely used markers for GAS exhibit limited sensitivity: MUC6 (51%) and HIK1083 (64%) (*50*), or PAX8 (68%) (*27*). By contrast, our dual-protein signature (PGC + DNMT1) achieves a diagnostic accuracy of 88% in the validation cohort. Together, these findings position the PGC-DNMT1 panel as a clinically deployable solution to the long-standing diagnostic ambiguity between GAS and HPV-CA.

PGC and DNMT1 each occupy distinct but complementary positions within the broader cancer landscape, lending therapeutic implications to our findings. PGC, a zymogen of pepsin C canonically produced by gastric chief cells, is an established serological and histological marker of gastric mucosal integrity, and its progressive loss represents a landmark early event in the Correa gastric carcinogenesis cascade (*51*). Notably, retained or elevated ectopic PGC expression has been significantly associated with more favorable outcomes in other epithelial tumor types, including mucinous ovarian carcinoma (*52*) and differentiated, ER-positive breast cancer subtypes, where it serves as an independent predictor of prolonged recurrence-free and overall survival (*51*), directly paralleling our observation that PGC-positive GAS patients exhibit a superior survival advantage. This cross-cancer consistency supports the paradigm of PGC as a generalizable indicator of highly differentiated, low-aggressiveness tumor phenotypes, whose depletion marks a malignant transition toward an aggressive, dedifferentiated state. At the genomic level, referencing the Genomic Data Commons (GDC) database (*53*), the PGC locus (mapped to chromosome 6p21.1) harbors a very low somatic mutation frequency in cervical cancer (*53*), consisting almost entirely of non-pathogenic passenger variants such as the single-nucleotide missense substitution A204T. This genomic intactness indicates that its dramatic expression divergence between GAS and HPV-CA is governed by lineage-specific transcriptional rewiring or epigenetic regulation rather than structural DNA alterations.

While PGC functions as an indicator for patient stratification in lineage-targeted strategies, DNMT1 represents a clinically actionable therapeutic target. Hypomethylating agents (HMAs) such as azacitidine and decitabine, which deplete DNMT1, are widely approved for myeloid malignancies (*54*) and have shown notable efficacy in advanced cervical cancer in a Phase II trial combining decitabine with cisplatin (*55*), despite encountering dose-limiting hematological toxicities. The elevated DNMT1 expression in HPV-CA reflects a classic state of virus-driven “epigenetic addiction,” mechanistically sustained by high-risk HPV E6-mediated p53 ubiquitination and degradation (*56*) and E7-mediated direct enzymatic activation via its CR3 zinc-finger domain (*57*), which renders these tumors vulnerable to DNMT1-targeted interventions. Consequently, a combined PGC-DNMT1 assessment yields highly complementary clinical metrics: PGC indexes the tumor differentiation state and lineage pedigree, while DNMT1 reflects the underlying viral etiology and delineates clear directions for targeted epigenetic and immunotherapeutic interventions.

To extend these findings to clinical prognosis, we further explored the clinical significance of PGC expression. While the majority of GAS cases are PGC-positive, our survival analysis revealed that the loss of PGC expression is a strong indicator of poor prognosis, possibly reflecting PGC’s role as a marker of cellular identity, whose absence could signify dedifferentiation or a shift toward a more aggressive phenotype. By integrating PGC status with standard clinicopathological variables, the resulting risk model achieved superior predictive performance compared with models based on clinical factors alone, enabling more effective patient stratification and providing a basis for personalized treatment planning. Together, these findings establish PGC as an independent prognostic factor and confirm its dual value as both a diagnostic hallmark and a standalone predictor of survival for GAS.

Several limitations of our study warrant mention. Although WGAN-GP helps address sample scarcity, synthetic data remain approximations of true biological distributions; despite our rigorous cross-validation, any potential for algorithmic bias requires cautious interpretation. Although the External Proteomic Cohort was independent, the proteomic sequencing for all samples was conducted centrally in a unified analytical workflow, intentionally designed to minimize technical batch effects during the biomarker discovery phase; future multi-center studies with decentralized sample processing and independent LC-MS analyses are warranted to fully confirm analytical reproducibility. Our mechanistic insights rely on proteomic associations; functional studies are necessary to determine whether these markers, particularly PGC, act as active oncogenic drivers or downstream effectors. The prognostic model was constrained by its retrospective nature and the limited cohort size of 95 patients; the lack of standardized intervention protocols for stratified risk groups also limits the immediate clinical application. Furthermore, the prognostic risk stratification relied on the empirical median of the training-derived Risk Score as the dichotomization threshold for Kaplan–Meier visualization. While data-driven cutoffs are common in exploratory analyses, this approach can inflate the apparent separation between risk groups. Finally, future investigations will explore the detectability of these markers in liquid biopsies such as serum or vaginal secretions, which would offer a promising avenue for non-invasive early screening.

Several coordinated steps are required to translate these findings into clinical practice. Independent prospective trials are needed to confirm the diagnostic accuracy of the PGC-DNMT1 panel across diverse populations and pathology practices. Mechanistic studies should clarify whether PGC loss is a driver of, or merely correlated with, aggressive GAS phenotypes, and whether DNMT1 expression can prospectively identify patients likely to benefit from hypomethylating or epigenetic therapies. Integration with existing pathology workflows, including automated staining quantification, would facilitate routine adoption. Together, these efforts would convert the current biomarker discovery into a clinically deployable, mechanistically grounded framework for the diagnosis and management of GAS.

Beyond its immediate clinical implications, the WEDGE framework illustrates how dual-stream graph learning combined with distribution-aware augmentation can extract interpretable biomarkers from rare-disease proteomics, offering a transferable approach for similar challenges in other small-cohort malignancies.

## MATERIALS AND METHODS

### Methods

This retrospective multi-center study was designed to discover and validate proteomic biomarkers for distinguishing gastric-type adenocarcinoma (GAS) from HPV-associated cervical adenocarcinoma (HPV-CA). The study comprised three prespecified components: (i) a proteomic discovery phase using bioinformatics analysis; (ii) a machine-learning driven biomarker selection phase using the WEDGE framework; and (iii) an independent validation phase combining immunohistochemistry and survival analysis.

### Patient characteristics

This study was approved by the Medical Ethics Committee of Zhejiang Cancer Hospital (Approval No. IRB-2025-76 (IIT), dated 23 January 2025). The requirement for informed consent was waived by the ethics committee because the study used anonymized archival formalin-fixed paraffin-embedded (FFPE) specimens and posed minimal risk to participants. The study was conducted in accordance with the Measures for Ethical Review of Life Science and Medical Research Involving Human Subjects (China), and the ICH-GCP guidelines. All patient data were de-identified prior to analysis. 407 FFPE cervical tissue specimens were retrospectively enrolled in this study, spanning the period from January 2015 to August 2024. Clinical biospecimens were derived from two distinct medical centers, where Zhejiang Cohort (n = 378) was collected from Zhejiang Cancer Hospital (Hangzhou, China) and designated as the internal discovery and development cohort, and Jiaxing Cohort (n = 29) was procured from Jiaxing Maternity and Child Health Care Hospital (Jiaxing, China) to serve as the geographically independent External Proteomic Cohort. The comprehensive baseline cohort structures integrated four distinct histological and physiological histotypes, comprising 125 GAS cases, 136 usual HPV-associated cervical adenocarcinoma (HPV-CA) cases, 16 rare non-GAS HPV-independent adenocarcinoma subtypes including 11 clear cell adenocarcinomas and 5 mesonephric adenocarcinomas, and 130 morphologically normal cervical tissues serving as control baselines. An independent IHC validation Cohort of 125 FFPE tissue specimens was assembled for immunohistochemistry-based validation of the WEDGE-identified biomarkers, comprising 50 GAS, 60 HPV-CA, and 15 morphologically normal cervical controls from Zhejiang Cancer Hospital. None of these specimens overlapped with the proteomic discovery or external proteomic cohorts. For the downstream prognostic survival analysis, a high-fidelity subset of 95 GAS patients from Zhejiang Cohort who exhibited comprehensive long-term post-operative follow-up profiles was utilized to construct the prognostic risk stratification model. To ensure strict phenotypic and histological purity, candidate participants across all comparative arms were dynamically screened based on rigorous, pre-specified eligibility pathways. Inclusion criteria for the malignant cohorts required histologically confirmed pure GAS or pure usual HPV-CA, which was independently evaluated and cross-verified by two senior gynecological pathologists based on standard morphological features combined with RNA-scope human papillomavirus detection results. Additionally, eligible cancer cases required the availability of sufficient tumor cellularity exceeding 60% within the archival FFPE tissue blocks, alongside complete and documented clinical, pathological, and demographic records. To establish an untainted physiological baseline and avoid the confounding molecular signatures frequently induced by tumor-adjacent field cancerization, the morphologically normal control samples were exclusively harvested from patients completely unaffected by cervical neoplastic diseases who underwent total hysterectomy for gynecological conditions, predominantly comprising uterine leiomyoma and adenomyosis. These control cervical tissues were structurally and histologically validated by the consensus pathologists to be entirely free of chronic inflammatory cervical lesions, squamous intraepithelial dysplasia, or any architectural atypia. Furthermore, specimens demonstrating mixed histopathological architectures, ambiguous lineage characteristics, or the co-existence of other synchronous or metachronous primary gynecological or systemic malignancies were entirely excluded from downstream computational modeling.

### Protein extraction, digestion and proteomic data acquisition

FFPE tissue blocks were serially sectioned at 10 μm thickness. Sections were dewaxed with xylenes (two changes, 10 min each), rinsed in absolute ethanol, and rehydrated through graded ethanol (100%, 90%, 75%; 5 min each) at 25°C. Samples were collected into tubes, and 100 mM Tris-HCl buffer (pH 10) was added for hydrolysis at 98°C for 1 hour with shaking at 800 rpm on a ThermoMixer. Protein digestion was performed using the filter-aided sample preparation method(*58*). Peptides were desalted using C18 tips (Pierce, Thermo Scientific, Rockford) per manufacturer’s instructions. Approximately 200 ng of peptides in 0.1% formic acid were analyzed on a Thermo Orbitrap Astral coupled to a Vanquish Neo LC (Thermo Fisher Scientific). Mobile phases consisted of 0.1% FA in water (A) and 80% ACN with 0.1% FA (B). Peptides were trapped and separated using trap and analytical columns as detailed in Data S1. Following ionization at 2.1 kV, data were acquired in DIA mode. Full scan spectra were collected at 240 k resolution (380-980 Da), while DIA scans used 2 m/z isolation windows across the same range. Maximum injection times were 5 ms and 3 ms for full and DIA scans, respectively. Additional parameters are provided in Data S1.

### Quality control and batch Design

Two pooled QC samples were prepared: (1) instrument_poolQC from 5 μL protein aliquots of 50 randomly selected cases for LC-MS stability monitoring; (2) method_poolQC from proteins pooled from another 50 random cases, inserted into each batch to monitor preparation stability. The 407 samples were processed in 8 batches. Each batch included 2 instrument_poolQCs, 1 method_poolQC, and 3 technical replicates of random samples. Batches 1-7 contained 50 samples each; batch 8 contained 57 samples.

### Data analysis and preprocessing

Raw data were searched against the UniProt human database (https://www.uniprot.org/; Feb. 2024) using Spectronaut 19 (Biognosys AG, Switzerland) in directDIA mode with trypsin as the digestion enzyme. Carbamidomethylation of cysteine was set as fixed modification; oxidation of methionine and N-terminal acetylation were variable modifications. FDR cutoffs were controlled at 1% at the precursor, peptide, and protein levels to ensure high confidence in identified and quantified proteins. Local normalization was applied, and major group quantities were calculated using MaxLFQ. Missing values were imputed from normal distributions using Perseus (v1.6.14.0; http://www.perseus-framework.org).

### Differential expression analysis

To comprehensively characterize the molecular landscape of cervical malignancies and delineate global proteomic reprogramming, downstream differential expression protein (DEP) analysis was systematically performed across the comprehensive evaluation cohort comprising 391 cases (excluding the 16 rare non-GAS variants from Zhejiang Cohort) in total, which included 114 GAS, 118 HPV-CA, and 130 controls from Zhejiang Cohort, alongside 11 GAS and 18 HPV-CA specimens from Jiaxing Cohort. These all-inclusive pairwise multi-group comparisons were systematically conducted to establish a holistic overview of the macro-level dysregulated protein profiles and were visually presented in fig. S2. Highly robust differentially expressed proteins were identified by executing an empirical Bayes moderated t-test within the ‘limma’ package (version 3.56.0) in the R statistical environment. To guarantee stringent control over the family-wise error rate and effectively minimize false positives inherent to high-throughput data processing, raw p-values were dynamically adjusted using the Benjamini-Hochberg (BH) false discovery rate (FDR) correction procedure, with global significance defined by an adjusted p-value < 0.05 combined with an absolute fold change > 1.5. Downstream functional characterization and systemic module interpretations based on these significantly differentially expressed proteins were executed via Gene Ontology (GO) and Kyoto Encyclopedia of Genes and Genomes (KEGG) pathway enrichment analyses utilizing the DAVID(*59*) bioinformatics platform (version 6.8), with the enrichment significance threshold locked at a Benjamini-Hochberg adjusted p-value < 0.05.

To prevent mathematical data leakage, informational contamination, and selection bias, the predictive WEDGE framework was entirely decoupled from the global population landscape. All core phases, including feature extraction, network topology construction, data augmentation, and biomarker ranking, were restricted strictly to network-interactive features identified via inner-loop gradient attributions within the isolated training cohort (n = 144).

### WEDGE framework for biomarker discovery

To enhance the generalization ability and interpretability required for biomarker discovery, we developed a novel computational framework called WEDGE (Wasserstein GAN-Enhanced Dual-stream Graph Convolutional Network for Expression analysis). This framework integrates data augmentation with graph-based deep learning to identify potential biomarkers from proteomic data. WEDGE consists of two core components: (1) a Wasserstein GAN with Gradient Penalty (WGAN-GP) module for proteomics data augmentation, and (2) a Dual-stream Graph Convolutional Network (GCN) that simultaneously processes protein-protein interaction (PPI) and gene regulatory network (GRN) information for disease classification and biomarker identification.

### WGAN-GP data augmentation module

A fundamental challenge in proteomics-based disease classification is the limited number of patient samples relative to the high dimensionality of protein expression features. This imbalance can lead to overfitting and poor generalization of machine learning models. To address this issue, we employed WGAN-GP to generate synthetic proteomic data that preserves the statistical properties of real samples while expanding the training dataset.

The WGAN-GP module consists of two neural networks: a Generator (G) and a Critic (D). The Generator learns to produce synthetic protein expression profiles from random noise vectors, while the Critic distinguishes between real and generated samples. Unlike traditional GANs, WGAN-GP uses the Wasserstein distance as the loss metric and incorporates a gradient penalty term to ensure stable training.

The Generator transforms a latent vector *z* ∈ *R*^100^ into a synthetic protein expression profile 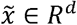, where *d* is the number of protein features. The Generator consists of four fully connected layers with LeakyReLU activation functions:

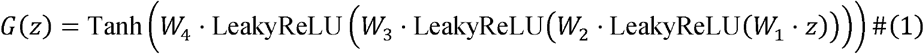

The layer dimensions are:100 → 250 → 500 → 1000 → *d*, where corresponds to the number of proteins.

The Tanh activation in the final layer ensures generated values fall within the normalized range [−1,1]. The Critic network mirrors the Generator structure in reverse, mapping protein expression profiles to scalar scores that estimate the Wasserstein distance:

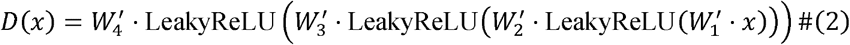

The layer dimensions are: *d* → 1000 → 500 → 250 → 1

The WGAN-GP training objective minimizes the Wasserstein distance with gradient penalty:

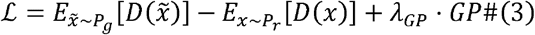

where *P*_*r*_ represents the real data distribution, *P*_*g*_ represents the generated data distribution, and *GP* is the gradient penalty term:

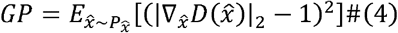

Here, 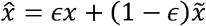 with ϵ ~ *U* (0,1) represents interpolated samples between real and generated data. We set *λ*_*GP*_ = 10 following standard practices. The training alternates between updating the Critic for *n*_*critic*_ = 3 iterations and updating the Generator once. Both networks are optimized using Adam optimizer with learning rate *α* 5 × 10 ^−5^ and momentum parameters *β*_1_ = 0.0, *β*_2_ = 0.9. Before training, protein expression data are normalized using z-score standardization computed on the training fold and applied to both training and validation folds.

To ensure robust model evaluation, we employ stratified 5-fold cross-validation. During this phase of model training and data augmentation parameter tuning, the models were optimized strictly using internal validation folds. For each fold, the WGAN-GP is trained independently on the training subset for 10,000 epochs with batch size of 16. To strictly prevent data leakage and ensure a robust evaluation, the WGAN-GP data augmentation was exclusively performed within the internal loop of the 5-fold cross-validation. For the total training cohort of 144 cases (73 GAS, 71 HPV-CA), each cross-validation fold partitioned approximately 115 samples (4/5) into the training subset and 29 samples (1/5) into the validation subset. The validation subset in each fold remained strictly isolated and unaltered, participating only in internal performance evaluation. The generator synthesized new data solely based on the training subset. With an augmentation factor of 5, the model generated five synthetic samples for each real training sample, yielding 575 synthetic samples per fold (290 GAS, 285 HPV-CA). These generated profiles were inverse-transformed to the original expression scale and merged with the 115 real training samples, establishing a balanced augmented training set of 690 samples per fold for downstream D-GCN training. Meanwhile, the initial 29 samples of the validation subset were utilized as an internal validation cohort to evaluate local classification performance and guide hyperparameter tuning. After which the optimized Fold 5 backbone was applied to the strictly held-out Internal Test Cohort (n = 88) and External Proteomic Cohort (n = 29) for unbiased performance assessment.

### Dual-Stream Graph Convolutional Network

The core architectural innovation of the WEDGE framework lies in its dual-stream graph convolutional network, which simultaneously models two distinct categories of physiological biological networks as structured heterogeneous graphs. Each individual patient specimen is mathematically represented as an independent heterogeneous graph, wherein nodes uniquely embody individual proteins or genes, and edges structurally encode validated biological relationships including protein-protein interactions and transcription factor-target regulatory linkages. Each patient sample is represented as a heterogeneous graph *G* = (*V, E*), where nodes *V* represent proteins/genes and edges *E* encode biological relationships (PPI or GRN network). For each patient, protein expression values serve as node features. Let *x*_*i*_ ∈ *R* denote the expression level of protein *i*. The node feature matrix is defined as *X* ∈ *R* ^*n* × 1^, where *n*, is the total number of proteins. Prior to entering the graph convolutional pipeline, the single-dimensional raw expression scalars are passed through a dedicated multi-channel node feature embedding layer configured as a learnable linear projection pipeline. To ensure a fair comparison between the two streams, the identical patient-specific protein expression matrix is fed to both the PPI and GRN branches. The embedding phase utilizes a module dictionary containing distinct linear transformation operators to project the raw features of both the mapped nodes from their initial one-dimensional space into a homogenized sixty-four-dimensional continuous latent representation space. This initial semantic alignment layer generates a dense initial hidden state matrix H^(0)^ ∈ R^n × 64^ thereby empowering the subsequent message-passing layers to capture rich, high-dimensional topological interactions and preventing structural information constriction.

Following the continuous latent feature projection, the structural topology of the heterogeneous graph is governed by two parallel edge tracking pathways designated as the PPI Flow and the GRN Flow. The protein-protein interaction network, defining the undirected PPI Flow, extracts functional and physical protein-protein association edges from the STRING database(*60*). The Protein-protein interaction edges *E*_*PPI*_ enforcing a stringent confidence score threshold equal to or exceeding 700 to filter out lower-probability experimental noise. These edges are undirected, reflecting bidirectional physical or functional associations between proteins. The adjacency matrix *A*_*PPI*_ is symmetrized:

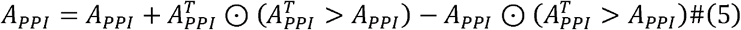

Conversely, the GRN network, defining the directed GRN Flow. The *E*_*GRN*_ are obtained validated transcription factor-target gene relationships from TRRUST(*61*) database to capture the unidirectional, causal regulatory influences driving downstream expression changes. These edges are directed, capturing the unidirectional regulatory influence. The adjacency matrix *A*_*GRN*_ preserves directionality. Both adjacency matrices are row-normalized. The D-GCN employs a heterogeneous convolutional architecture to process PPI and GRN networks in parallel. Each stream consists of two graph convolutional layers with residual connections and graph normalization.

For the convolution layer 1, the convolution aggregates neighborhood information separately for each edge type:

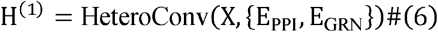

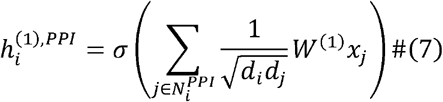

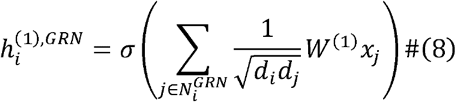

where 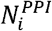 and 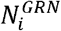 denote the neighbors of node *i* in PPI and GRN networks respectively, *d*_*i*_ is the degree of node *i, W* ^(1)^ is a learnable weight matrix, and *σ* is the ReLU activation function. After convolution, we apply graph normalization to stabilize training:

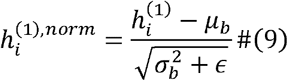

where *μ*_*b*_ and 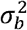 are the mean and variance computed over nodes in the same batch assignment. The residual connection helps preserve information from earlier layers and facilitates gradient flow during backpropagation.

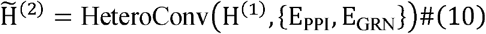

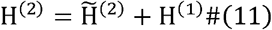

To obtain graph-level representations from node-level features, we employ both mean and max pooling:

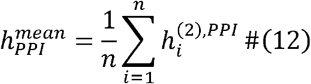

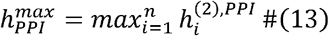

The pooled features are concatenated:

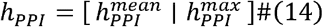

The same pooling strategy is applied to the GRN stream to obtain *h*_*GRN*_. Each stream has a dedicated multi-layer perceptron (MLP) classifier:

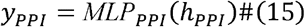

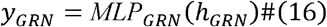

To integrate predictions from both streams, we employ a learnable attention mechanism:

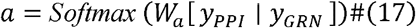

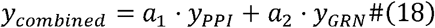

Where 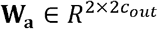 is a learnable weight matrix, and *a*_1,_ *a*_2_ are attention weights summing to 1.

The D-GCN was optimized with the Adam optimizer (initial learning rate *α* = 1 × 10 ^−3^ weight decay *λ* = 1 × 10 ^−5^ under mixed-precision (16-bit) training. We employed a learning-rate scheduler that reduced the learning rate on plateau (factor 0.5, patience 20), together with EarlyStopping (patience 100, min Δ = 1 × 10 ^−4^) monitoring validation loss, and a minimum training of 1,000 epochs (maximum 2,000 epochs). For visualization in fig. S3, training trajectories were uniformly truncated to the first 6,000 steps across all five cross-validation folds to enable visual comparison.

All models were implemented using PyTorch 2.1.0, PyTorch Geometric 2.6.1, and PyTorch Lightning 2.4.0. Training was conducted on NVIDIA GPU 2080ti with mixed-precision (16-bit) training to accelerate computation.

### Integrated Gradients for Feature Attribution

The interpretability of WEDGE is explicitly realized by mapping the learned attention weights back to specific protein nodes within the PPI and GRN networks. This attribution technique quantifies the contribution of each protein’s expression to the model’s prediction by integrating gradients along a path from a baseline input to the actual input, identifying key drivers like PGC rather than numerical outputs.

For a given patient sample with protein expression *x* = (*x*_1_, *x*_2_, … *x*_*n*_), we define a baseline *x′* = 0 representing a reference state with zero expression. The integrated gradient for protein *i* is computed as:

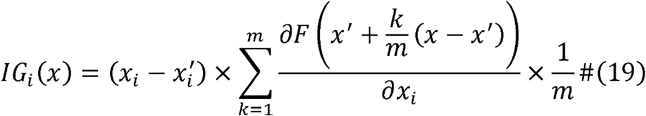

where *F*(*x*) represents the WEDGE model’s output logit for the target class, and *m* = 50 is the number of interpolation steps. The importance score for protein *i* is the absolute value of *IG*_*i*_ (x). For both attribution methods, we compute importance scores for each patient across the internal validation folds and aggregate them:

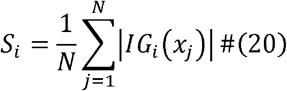

where *N* is the number of samples and *S*_*i*_ represents the average importance of protein *i*. Proteins are ranked by their aggregated scores, and top-ranked proteins are considered candidate biomarkers. This feature ranking process was completely nested within the cross-validation framework of the training Cohort, ensuring strict isolation from the Internal Test Cohort and External Proteomic Cohort.

### Consensus-based Feature Selection

For the combination selection stage, in order to identify a parsimonious yet robust diagnostic signature, we implemented a consensus-based selection protocol. We systematically evaluated 4000 possible protein combinations across multiple panel sizes (k = 3, 4, 5, 6, 7, 8, 9, 10). Because the total number of combinations increases sharply with 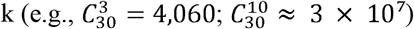, we capped each panel size at 4,000 randomly sampled combinations to ensure uniform sampling across all panel dimensions and to avoid selection bias toward smaller panel sizes. For each size, the metric was defined as the mean AUC across the validation folds.

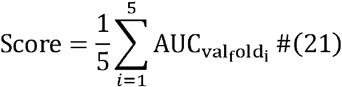

To avoid the selection of biased markers that perform exceptionally well in only one cohort or are carried by a single dominant feature, we quantified the recurrence frequency of each protein across the top 50 combinations of each panel size. Proteins that consistently appeared with the highest frequency across all dimensions (PGC and DNMT1) were prioritized for the final 2-protein diagnostic panel.

### Performance evaluation

To assess the quality of WGAN-GP generated samples, we compute the Fréchet Distance (FD)(*24*) between real and synthetic protein expression distributions. We computed FD directly in the original n-dimensional protein space, where n is the number of quantified proteins.

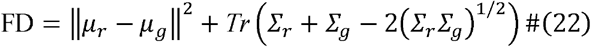

Where *μ*_*r*_ and *μ*_*g*_ represent the mean vectors of real and generated protein expression distributions, respectively, *Σ*_*r*_ and *Σ*_*g*_ represent their covariance matrices, and Tr(·) denotes the trace operator. Lower FD scores indicate higher quality synthetic data that better approximates the real data distribution. As a baseline, we compare against randomly generated samples to validate that WGAN-GP provides meaningful augmentation.

For the final evaluation, the finalized model architecture and the selected two-protein panel (PGC and DNMT1) were evaluated exclusively on the strictly sequestered Internal Test Cohort (n = 88) and the independent External Proteomic Cohort (n = 29) to assess true generalization performance. Model performance is evaluated using multiple metrics derived from the confusion matrix:

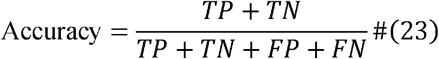

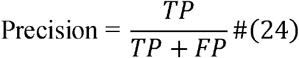

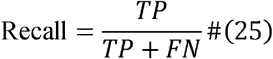

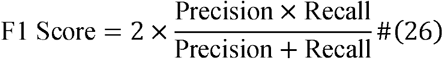

where *TP, TN, FP*, and *FN* denote true positives, true negatives, false positives, and false negatives, respectively. We plot Receiver Operating Characteristic (ROC) curves by varying the classification threshold and computing the True Positive Rate (TPR) and False Positive Rate (FPR):

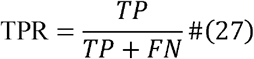

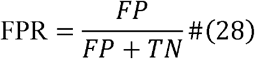

The Area Under the ROC Curve (AUC) serves as a threshold-independent metric of model discrimination ability.

### Immunohistochemistry and interpretation

To evaluate the clinical translatability and diagnostic scalability of the WEDGE-identified biomarkers within routine pathological workflows, we assembled an independent IHC validation cohort. Specimens were fixed in 10% neutral buffered formalin and embedded in paraffin. To address potential intra-tumoral heterogeneity and ensure diagnostic clarity, our study strictly included cases of pure gastric-type adenocarcinoma (GAS) or pure HPV-associated adenocarcinoma (HPV-CA), explicitly excluding any tumors with mixed-type histological features. For each case, 1 to 3 representative paraffin blocks were selected to prepare 3 to 5 μm thick sections. Immunohistochemical (IHC) testing was performed using the EnVision two-step method. Sections were deparaffinized in xylene, rehydrated through graded ethanol, and subjected to heat-induced epitope retrieval (HIER) in a pressure cooker. Tissue sections were subsequently incubated with validated primary antibodies directed against PGC and DNMT1 under optimized dilution profiles using positive tissue matrices as continuous positive benchmarks alongside parallel antibody-omitted negative controls. Endogenous peroxidase activity was blocked with 3% hydrogen peroxide. The following primary antibodies were used: PGC (Proteintech, 28532-1-AP), DNMT1 (Proteintech, 24206-1-AP). Sections were incubated with primary antibodies, followed by the secondary antibody (horseradish peroxidase-conjugated EnVision + System-HRP). Staining was visualized using 3,3’-diaminobenzidine (DAB) and counterstained with hematoxylin. Appropriate positive and negative controls were run concurrently for all antibodies.

All stained immunohistochemical slides were systematically evaluated via a consensus-driven multi-head microscopy configuration by senior gynecological pathologists who maintained an absolute blind state regarding the patient classifications, pathological tumor staging, and clinical outcomes. Cases with IHC staining for PGC and DNMT1 were included in the machine learning classification analysis. For each biomarker, two quantitative features were extracted: staining intensity (Strength) and positive cell proportion (Percentage), resulting in four features in total. The feature combinations were systematically evaluated: (1) PGC and DNMT1 combined; (2) PGC alone; and (3) DNMT1 alone. The dataset was randomly partitioned into training (70%) and testing (30%) sets using stratified sampling. An XGBoost classifier was employed for binary classification between GAS and HPV-CA. All analyses were performed using Python with scikit-learn (version 1.7.1) for model evaluation and XGBoost (version 3.0.4) for classification. To mathematically evaluate and compare the statistical distributions of these continuous pathological scores across the distinct clinical groups, historical baseline statistical significance testing was performed utilizing GraphPad Prism software (version 10.1.2). Multi-group cohort variance evaluations simultaneously encompassing the GAS, HPV-CA, and normal control groups were analyzed via a one-way analysis of variance (ANOVA) followed by Tukey’s multiple comparisons test, with statistical significance strictly defined by a multi-scale adjusted p-value cutoff (*P < 0.05, ** P < 0.01, and ***P < 0.001).

### Prognosis model construction

All 95 GAS participants with complete follow-up data and IHC staining for PGC were included in the prognostic model analysis (median follow-up: 38 months; range: 2-96 months). The primary endpoint was overall survival (OS), defined as the time from surgery to death from any cause or last follow-up. The 95 patients were randomly partitioned into a Training Cohort (n = 63, 66%) and a Test Cohort (n = 32, 34%) using stratified sampling on the event indicator (death vs. censored). Initially, univariate Cox proportional hazards regression was performed to evaluate the association between individual clinicopathological factors, PGC expression status (Negative vs. Positive), and overall survival (OS). To integrate multiple risk factors into a unified predictive tool, we employed a Random Survival Forest (RSF) algorithm. The forest ensemble was trained on the continuous and categorical clinicopathological variables alongside the dichotomized PGC expression profiles to generate a continuous prognostic Risk Score for each individual patient via out-of-bag feature permutation tracking, automatically stratifying the clinical cohort into High-Risk and Low-Risk prognostic sub-populations utilizing the empirical median Risk Score as the absolute mathematical separation threshold. Variables demonstrating statistical significance (P<0.05) in the univariate analysis were further incorporated into a multivariate Cox proportional hazards regression model to identify independent prognostic factors and calculate adjusted hazard ratios (HRs) with 95% confidence intervals. Model performance was evaluated using Kaplan-Meier survival curves with log-rank tests to compare survival differences between risk groups, receiver operating characteristic (ROC) curves with the area under the curve (AUC) to assess predictive accuracy, and Harrell’s concordance index (C-index) to evaluate discriminative ability. All statistical analyses were performed using R software (version 4.3.2, RStudio version 2023.06.1) with the survival package (version 3.6.4) and survminer (version 0.4.9) packages for Cox modeling, randomForestSRC (version 3.4.1) for risk score generation, and timeROC (version 0.4) for predictive performance evaluation. p < 0.05 was considered statistically significant.

## Data Availability

All data and code produced in this work are available online. The mass spectrometry proteomics data are deposited in the ProteomeXchange Consortium via the iProX repository (identifier: PXD074127 / IPX0013995000) and will be released publicly upon manuscript publication at https://www.iprox.cn/page/PSV023.html;?url=1782219624318PdxleQ1V. Source code is openly accessible at https://github.com/HuangHan-LabAccount/WEDGE-for-biomarker-discovery.

## Data availability

The mass spectrometry proteomics data have been deposited to the ProteomeXchange Consortium via the iProX partner repository with the dataset identifiers PXD074127 (IPX0013995000).

## Code availability

Code is available at https://github.com/HuangHan-LabAccount/WEDGE-for-biomarker-discovery

## Acknowledgments

We thank Wangang Gong for performing the immunohistochemistry assays, Xianhua Fang for the expert pathological evaluation of the IHC results, and Zhan Zhou for advice on the methodological architecture of the study.

## Author contributions

Conceptualization: T.Z., H.H.

Methodology: T.Z., H.H., M.N.

Software: H.H.

Validation: Y.X., X.F.

Formal analysis: H.H., M.N.

Investigation: T.Z., H.H., M.N., Y.X.

Resources: T.Z., X.T., X.F.

Data curation: H.H., M.N.

Writing original draft: T.Z., H.H.

Writing, review & editing: Z.Y., W.L., Y.L., C.Y., Z.S., J.W.

Visualization: H.H.

Supervision: Q.W., J.W., H.T.

Project administration: T.Z.

Funding acquisition: Q.W., T.Z., J.W.

## Data, code, and materials availability

Formalin-fixed paraffin-embedded tissue specimens used in this study were obtained from the pathology archives of Zhejiang Cancer Hospital and Jiaxing Maternity and Child Health Care Hospital. Commercial primary antibodies (anti-PGC, Proteintech 28532-1-AP; anti-DNMT1, Proteintech 24206-1-AP) were obtained from commercial suppliers and are publicly available for purchase. The mass spectrometry proteomics data have been deposited to the ProteomeXchange Consortium via the iProX partner repository with the dataset identifier PXD074127 (IPX0013995000).The source code is publicly available at https://github.com/HuangHan-LabAccount/WEDGE-for-biomarker-discovery for community access.

## Supplementary Materials

**Fig. S1.**
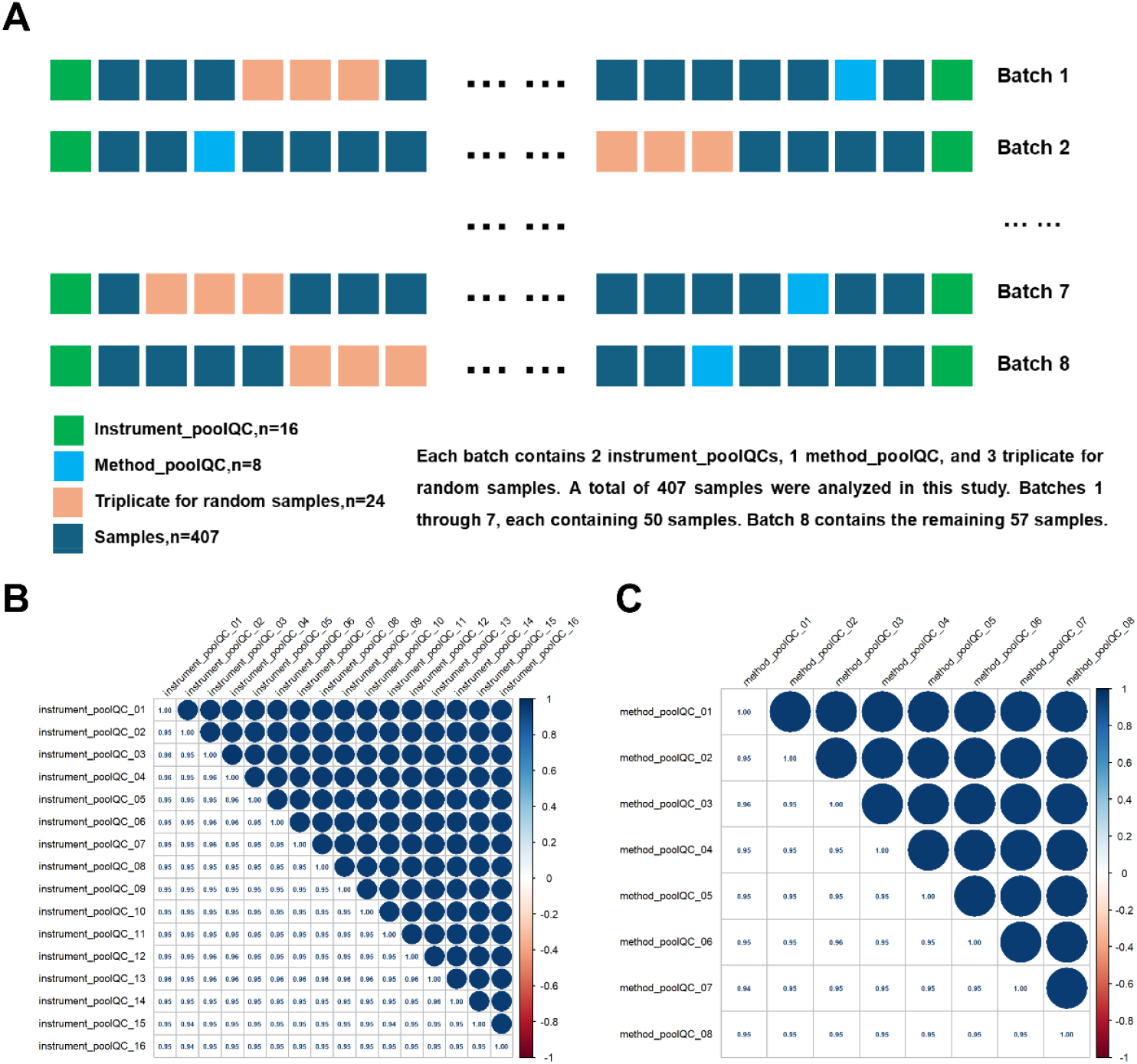
Multi-batch quality control of the LC-MS/MS proteomic workflow. (**A**)Schematic of the batch design. Eight continuous technical blocks were used to process 407 samples in total. Each batch contained 2 instrument-pool QCs (n = 16 total), 1 method-pool QC (n = 8 total), 3 triplicates of random samples (n = 24 total), and the remaining study samples. Batches 1-7 each contained 50 samples, and batch 8 contained the remaining 57 samples. (**B**) Cross-batch Pearson correlation matrix for the 16 instrument-pool QC samples. (**C**) Cross-batch Pearson correlation matrix for the 8 method-pool QC samples.

**Fig. S2.**
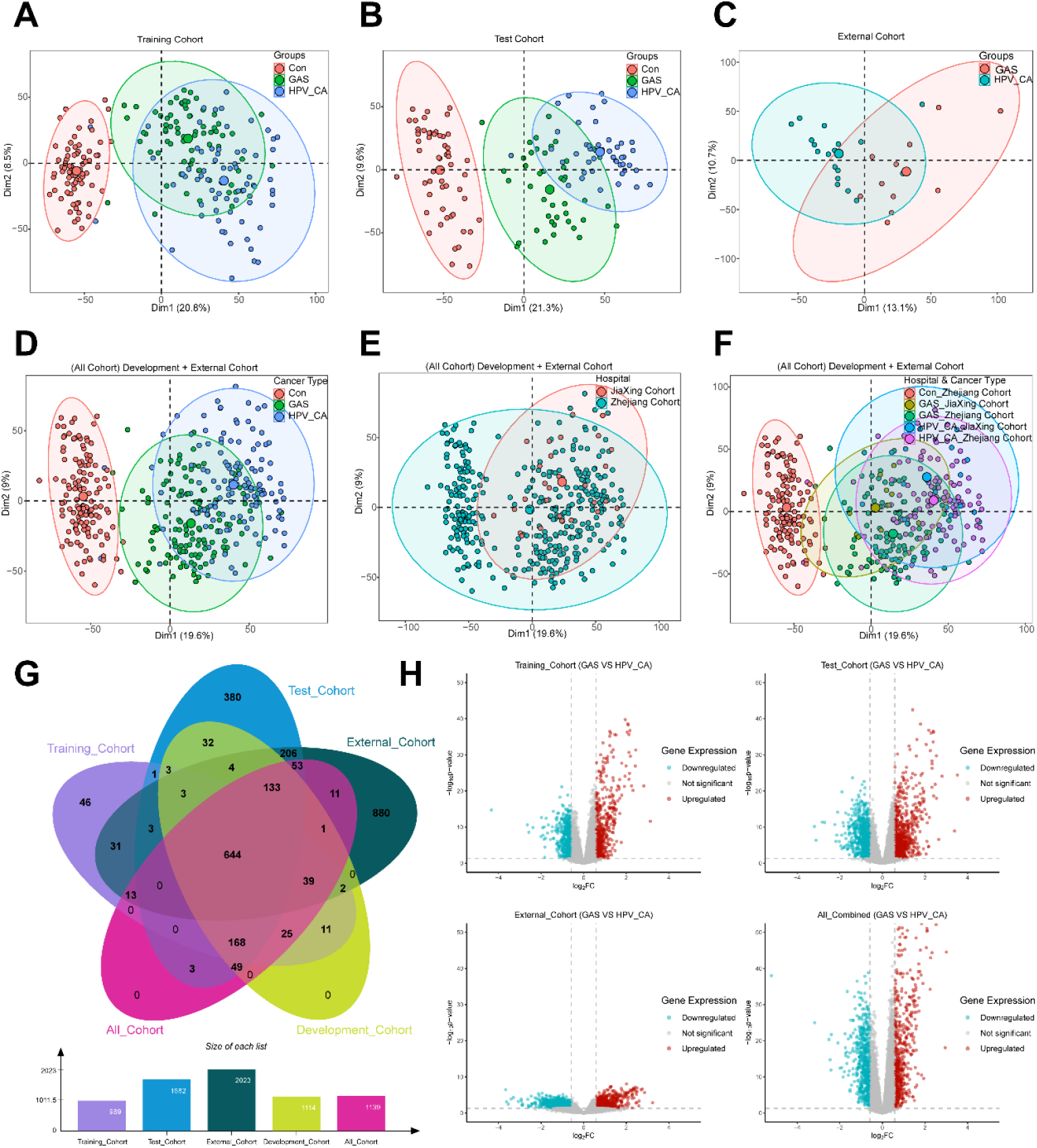
PCA and DEP analysis of proteomic cohorts. (**A-C**)PCA of proteomic profiles split by cohort: Training Cohort (A), Test Cohort (B) and External Cohort (C), coloured by sample group (Con / GAS / HPV-CA). (**D-F**) PCA of the combined Development + External Cohort coloured by cancer type (D), hospital (E) and hospital-by-cancer-type (F). GAS and HPV-CA samples overlap substantially, indicating that traditional linear dimensionality reduction cannot resolve the biochemical differences between the two adenocarcinoma subtypes. (**G**) Venn diagram showing the overlap of differentially expressed proteins (DEPs) across the Training, Test, External, Development and All-Combined cohorts. (**H**) Volcano plots of the Training, Test, External and All-Combined cohorts (GAS vs. HPV-CA).

**Fig. S3.**
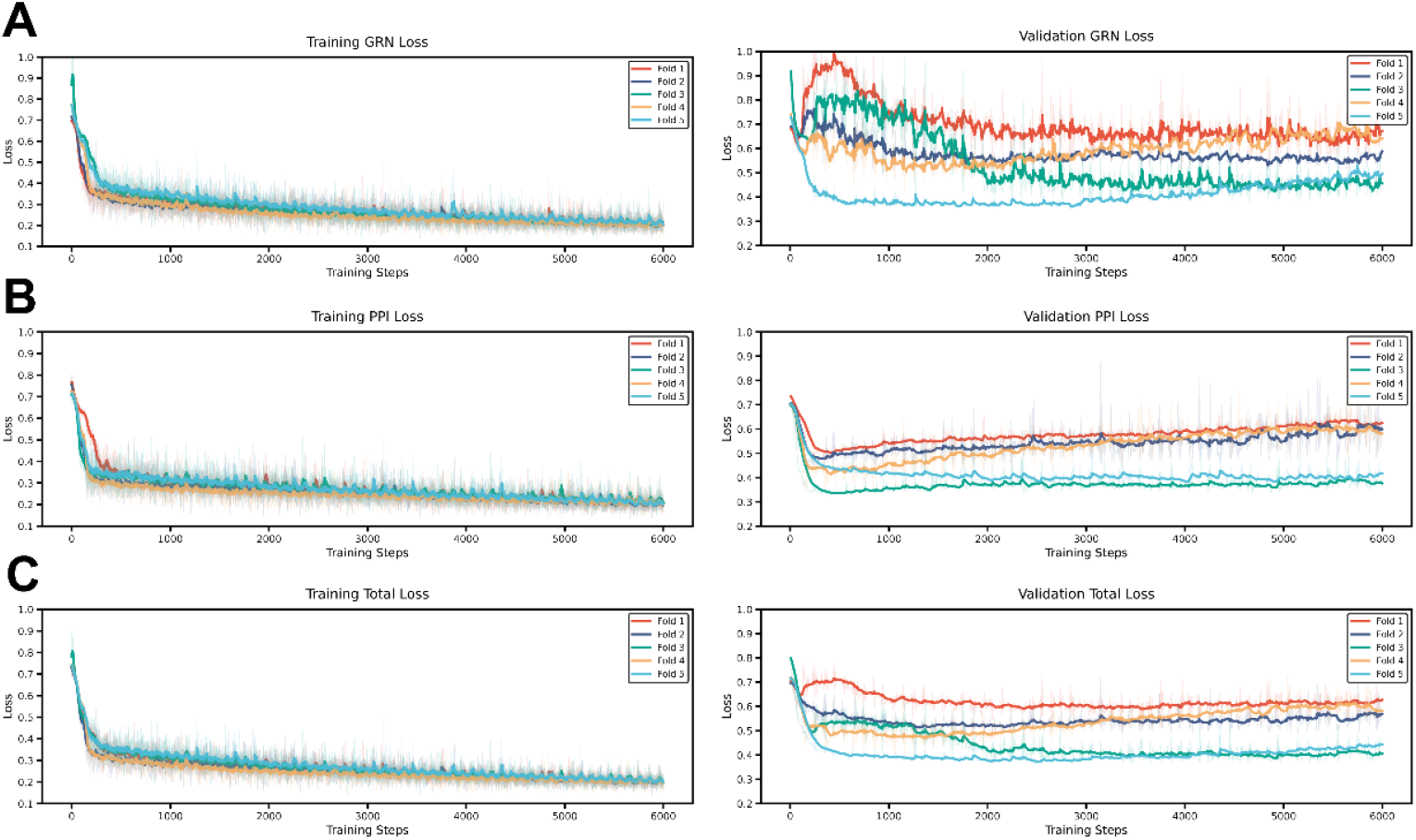
Training trajectories of the dual-stream graph neural network. (**A**) Training (left) and validation (right) loss curves for the Gene Regulatory Network (GRN) stream. (**B**) Training (left) and validation (right) loss curves for the Protein-Protein Interaction (PPI) stream. (**C**) Training (left) and validation (right) Total Loss curves. All curves are plotted across 6000 training steps for the five folds.

**Fig. S4.**
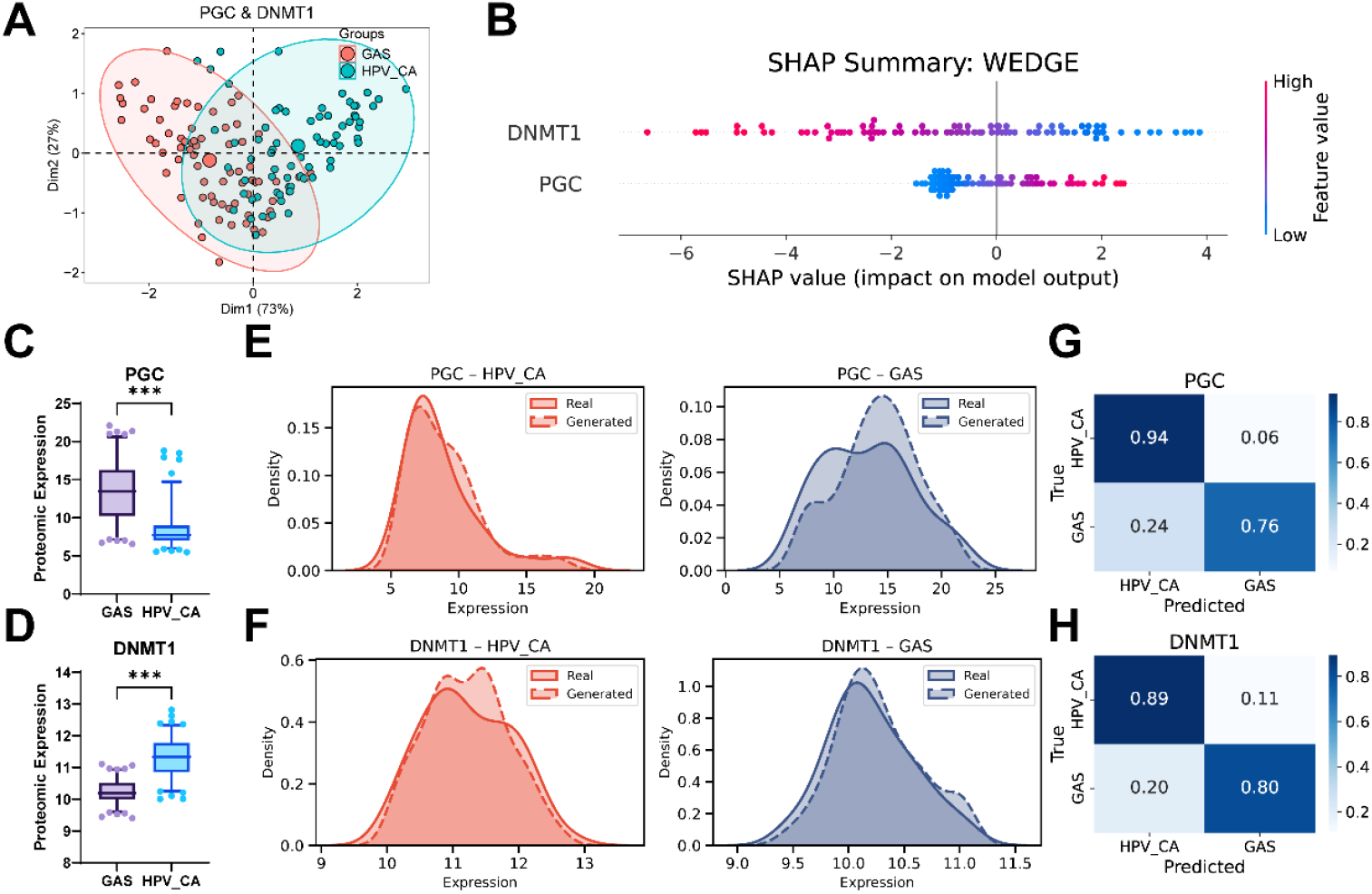
Detailed evaluation of the WEDGE-identified two-protein biomarker panel (PGC and DNMT1) (**A**) Unsupervised PCA using only the expression values of PGC and DNMT1 separates GAS and HPV_CA into distinct clusters, reproducing the separation pattern observed with the full proteome. (**B**) SHAP summary plot for the WEDGE model showing the relative contributions and directions of PGC and DNMT1 on the model output. (**C, D**) Proteomic expression of PGC (C) and DNMT1 (D) in GAS versus HPV_CA (***p < 0.001). (**E, F**) Probability density distributions demonstrating latent-distribution matching, where WGAN-GP-generated profiles (dashed lines) closely mirror the empirical distributions of the real patient cohorts (solid lines) across both HPV_CA and GAS classes for PGC (E) and DNMT1 (F). (**G, H**) Confusion matrices evaluating single-marker classification performance for PGC (G) and DNMT1 (H).

**Fig. S5.**
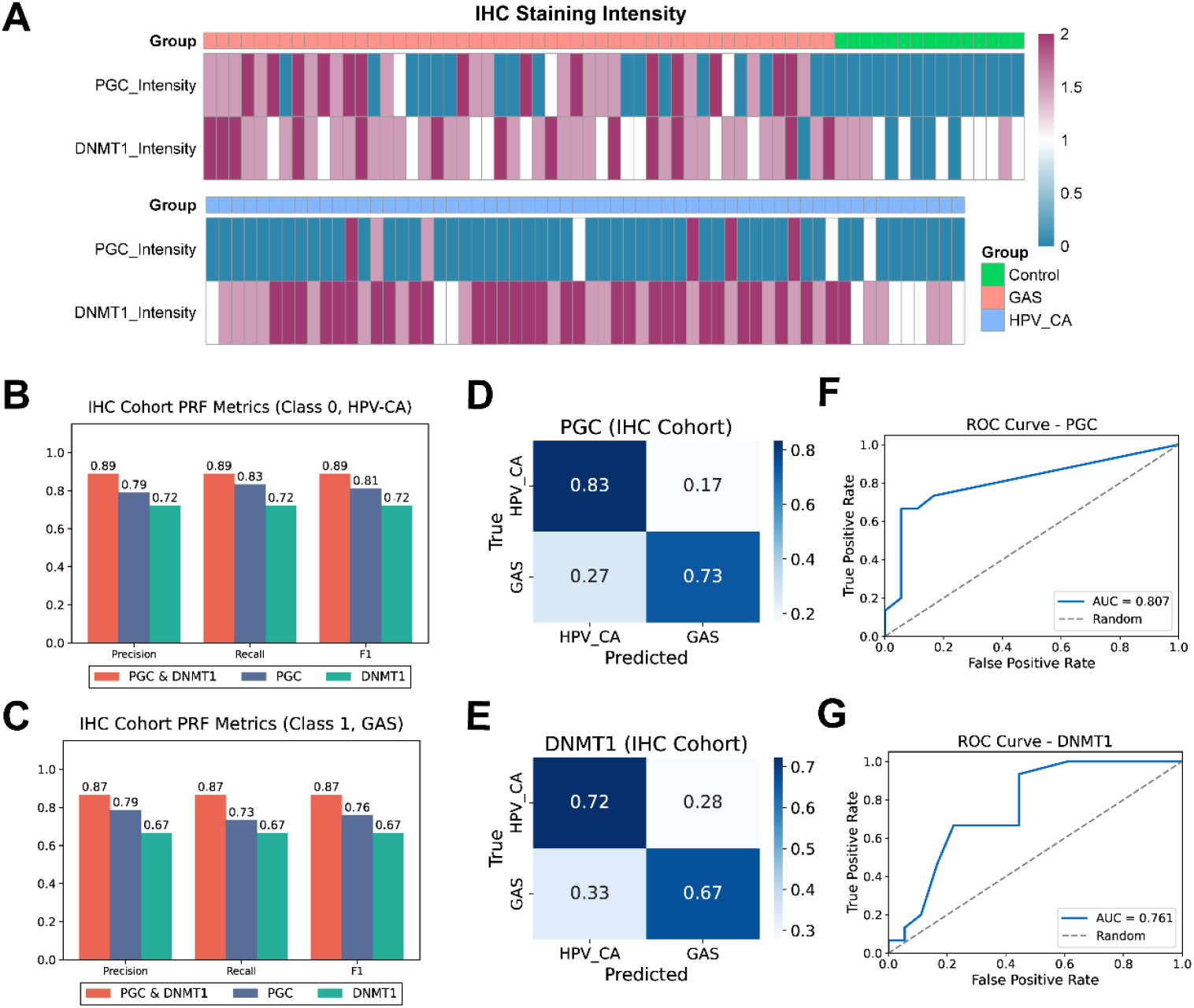
Immunohistochemistry (IHC) validation of the PGC and DNMT1 biomarkers. (**A**) Heatmaps of IHC staining intensity for PGC and DNMT1 across the three groups (Control, GAS, HPV_CA), demonstrating the differential expression patterns of the two biomarkers in routine pathological specimens. (**B, C**) Precision, recall and F1-score metrics of the combined panel (PGC and DNMT1) versus the single biomarkers for Class 0 HPV_CA (B) and Class 1 GAS (C). (**D, E**) Confusion matrices demonstrating the diagnostic classification performance in the IHC validation cohort for PGC (D) alone and DNMT1 (E) alone. (**F, G**) Receiver Operating Characteristic (ROC) curves showcasing the discriminatory capacity of the single-biomarker models, including PGC (F) alone and DNMT1 (G) alone. These are enlarged views of the single-marker ROC curves originally shown in Fig. 6E to facilitate inspection of the AUC values and the shape of each curve.

**Fig. S6.**
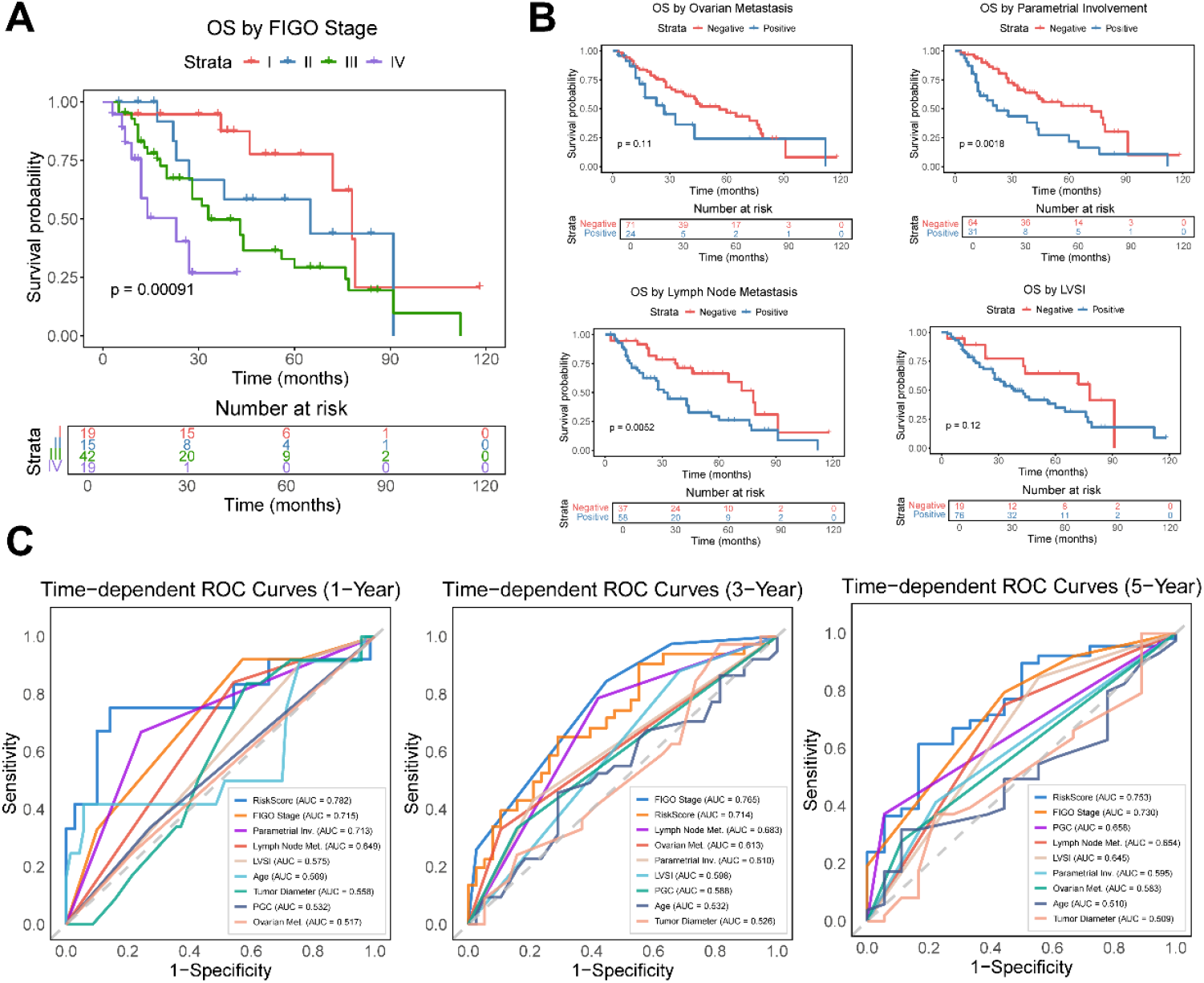
Prognostic analyses of clinicopathological parameters. (**A**) Kaplan-Meier overall survival (OS) curves stratified by FIGO stage, with the number-at-risk table shown beneath. (**B**) Kaplan-Meier OS curves for four additional standard clinicopathological variables: ovarian metastasis, parametrial involvement, lymph-node metastasis and LVSI, each with its number-at-risk table. (**C**) Time-dependent ROC curves at the 1-, 3- and 5-year horizons comparing the Risk Score with FIGO stage, parametrial involvement, lymph-node metastasis, LVSI, age, PGC, tumour diameter and ovarian metastasis.

**Data S1. LC-MS proteomic batch and quality-control overview across the 407-sample cohort**

Spreadsheet containing the full batch and quality-control (QC) overview for the LC-MS/MS proteomic profiling of all 407 cervical tissue samples. The remaining sheets document the column and liquid-chromatography parameters, the Orbitrap Astral acquisition settings.

**Data S2. WGAN-GP-generated synthetic proteomic profiles**

This file contains the synthetic proteomic profiles generated from the optimal cross-validation fold (Fold 5, which achieved the lowest validation loss and Fréchet Distance) (n = 575, including two class-specific subsets for GAS and HPV-CA). Due to file size limitations, the generated data for other training folds are available from the corresponding authors upon reasonable request.

**Data S3. Network-importance scores for the PPI and GRN candidate proteins underlying WEDGE**

**Data S4. IHC intensity scores for PGC and DNMT1 in the independent validation cohort**

